# Identifying potential risk genes and pathways for neuropsychiatric and substance use disorders using intermediate molecular mediator information

**DOI:** 10.1101/2023.03.15.23287330

**Authors:** Huseyin Gedik, Tan Hoang Nguyen, Roseann E. Peterson, Christos Chatzinakos, Brien P. Riley, Vladimir I. Vladimirov, Silviu-Alin Bacanu

**Affiliations:** Integrative Life Sciences, Virginia Institute of Psychiatric and Behavioral Genetics, Virginia Commonwealth University, Richmond, VA, USA; Department of Psychiatry, Virginia Institute for Psychiatric and Behavioral Genetics, Virginia Commonwealth University, Richmond, VA, USA; Institute for Genomics in Health, SUNY Downstate Health Sciences University, Brooklyn, NY, USA; Department of Psychiatry and Behavioral Sciences, SUNY Downstate Health Sciences University, Brooklyn, NY, USA; Stanley Center for Psychiatric Research, Broad Institute of MIT and Harvard, Cambridge, MA, USA; Department of Psychiatry, McLean Hospital, Harvard Medical School, Belmont, MA, USA; Department of Psychiatry, College of Medicine-Phoenix, University of Arizona, Phoenix, AZ, USA

**Keywords:** eQTL, pQTL, mQTL, Mendelian randomization, Alcohol Use Disorder, Bipolar Disorder, Schizophrenia

## Abstract

Neuropsychiatric and substance use disorders (NPSUD) have a complex etiology that includes environmental and polygenic risk factors with significant cross-trait rG. Genome Wide Association Studies (GWAS) of NPSUD yield numerous association signals. However, for most of these regions, we do not yet have a firm understanding of either the specific risk variants or the effects of these variants. Post-GWAS methods allow researchers to use GWAS summary statistics and functional genomics data to infer the likely molecular mediators (transcript, protein and methylation abundances) for the effect of variants on disorders. One group of post-GWAS approaches is commonly referred to as transcriptome/proteome/methylome wide association studies, which are abbreviated as T/P/MWAS (or collectively as XWAS). Since these approaches use biological mediators, the multiple testing burden is reduced to the number of genes (∼20,000) instead of millions GWAS SNPs leading to increased signal detection. In this work, our aim is to uncover likely risk genes for NPSUD by performing XWAS analyses in two tissues – blood and brain. Firstly, XWAS using the Summary-data based Mendelian Randomization (SMR), which takes GWAS summary statistics, reference xQTL data and a reference LD panel as inputs, was conducted to identify putative causal risk genes. Second, given the large comorbidities among NPSUD and the shared cis-xQTLs between blood and brain, we improved XWAS signal detection in NPSUD for underpowered analyses by performing joint concordance analyses between XWAS results i) across the two tissues and ii) across NPSUD. All XWAS signals i) were adjusted for HEIDI (non-causality) p-values and ii) used to test for pathway enrichment. The results suggest that there were widely shared gene/protein signals within the Major Histocompatibility (MHC) region on chromosome 6 (*BTN3A2* and *C4A)* and elsewhere in the genome (*RERE, FURIN, ZDHHC5* and *NEK4*). The identification of putative molecular genes and pathways underlying risk may offer new targets for therapeutic development. Some of our analyses’ more immediate actionable signals might relate to vitamins, i.e., i) in *KYAT3* (a part of the kynurenine pathway with vitamin B6 as a cofactor) for post-traumatic stress disorder and ii) omega-3 and vitamin D pathways for bipolar disorder.

## 1. Introduction

Genome-wide association studies (GWAS) have identified numerous loci associated with neuropsychiatric and substance use disorders (NPSUD), supporting the high polygenicity of NPSUD. Furthermore, NPSUD’s risk loci have not been fully discovered (Owen and Williams, 2021). For instance, the largest GWAS for schizophrenia (SCZ) found 287 independent loci and estimates that the common variants explain only 24% of the phenotypic variance (Trubetskoy et al., 2022). Similarly, other NPSUD GWAS yield large numbers of genome-wide significant signals (Howard et al., 2019; Nievergelt et al., 2019; Sanchez-Roige et al., 2019; Mullins et al., 2021) capturing a statistically significant but small proportion of the phenotypic variance. Since most genome-wide significant signals reside in non-protein coding genomic regions (Edwards et al., 2013), the interpretation of these GWAS findings are not straightforward. Performing post-GWAS analyses, that infer associations between genes or molecular pathways and traits, could significantly advance our understanding of these GWAS signals.

Associated variants are thought to influence risk through altered gene regulation, for example, via epigenetic changes, yielding changes in RNA levels or protein abundance. Molecular quantitative loci mapping studies empirically support this assumption. They found that expression quantitative trait loci (eQTL) (Ongen et al., 2017), protein QTL (pQTL) (Robins et al., 2021) and methylation QTL (mQTL) (Hannon et al., 2016) colocalize with disease-associated loci. However, while there are many well-powered GWAS scans in NPSUD (Pardiñas et al., 2018; Demontis et al., 2019; Grove et al., 2019; Howard et al., 2019; Nievergelt et al., 2019; Watson et al., 2019; Johnson et al., 2020; Polimanti et al., 2020; Mullins et al., 2021; Trubetskoy et al., 2022), none of these studies directly assayed the transcriptome, proteome or methylome for their cohorts.

However, researchers found ways around this assessment limitation in GWAS cohorts. They formed large reference molecular e/p/mQTL (henceforth denoted as xQTL) datasets readily available for blood and brain (Sun et al., 2018; van der Wijst et al., 2020; Ferkingstad et al., 2021; Võsa et al., 2021; Yang et al., 2021a; Zhang et al., 2021). Researchers have developed methods to integrate these molecular xQTL data and GWAS summary statistics to impute the association between phenotypes and molecular mediators (transcriptome, proteome and methylome). Such analyses are widely referred to as transcriptome-wide association studies (TWAS), proteome-wide association studies (PWAS) and methylome-wide association studies (MWAS) (Gamazon et al., 2015; Gusev et al., 2016; Zhu et al., 2016; Barbeira et al., 2018, 2019; Hu et al., 2019; Nagpal et al., 2019; Bae et al., 2021) – henceforth collectively referred to as XWAS. Moreover, since they directly model relevant biological mediators, these approaches could identify putatively causal genes but they fail to eliminate pleiotropy (Wainberg et al., 2019). Until recently, XWAS analyses of NPSUD were mostly TWAS (Zhu et al., 2016; Niu et al., 2019; Hammerschlag et al., 2020; Kapoor et al., 2021). However, PWAS is also increasing in number with the expanding pQTL reference data in brain (Wingo et al., 2021, 2022). Besides eQTL and pQTL, mQTL has also been investigated as a risk factor (Perzel Mandell et al., 2021; Shen et al., 2022). Recently, MWAS also yielded significant genes for NPSUD (Sugawara et al., 2018; Aberg et al., 2020; Howard et al., 2022).

Changes in gross anatomical and cell type specific phenotypes have been observed for NPSUD and associated risk alleles via *in vitro* and postmortem studies (Brennand et al., 2012; Schrode et al., 2019; Zhang et al., 2020). Functional genomic profiles differ by cell type (Marstrand and Storey, 2014; Buenrostro et al., 2015) and cell type composition differs across brain regions (Wang et al., 2018). In addition, different neuronal cell types have different functional profiles and different distributions across regions (Kelley et al., 2018). Genetic variants contributing to the heritability of certain NPSUD were enriched in cis-regulatory elements that are specific to GABAergic and glutamatergic neurons (Sanchez-Priego et al., 2022). Thus, integrating cell type specific xQTL with GWAS findings is very promising. However, due to expense and other factors, sample sizes for functional profiles in specific cell types across brain regions are still small (Spaethling et al., 2017; Bryois et al., 2022) and limited in detection power. Consequently, most functional genomics data available to support xQTL mapping studies come from bulk brain tissue (Consortium, 2020) rather than single cell (Bryois et al., 2022) or sorted cell types (Aygün et al., 2021), and despite their limited cellular resolution, the use of bulk tissue and meta-analysis across tissues are currently still more powerful.

To conduct XWAS, two common approaches, TWAS (Gusev et al., 2016) and PrediXcan (Gamazon et al., 2015) have been used, with both requiring pre-computing of SNP weights from xQTL data sets. To avoid LD confounding, these XWAS tools also require a subsequent fine mapping step – e.g., TWAS-FOCUS (Mancuso et al., 2019). In contrast, Mendelian Randomization (MR) based methods (Zhu et al., 2016; Yuan et al., 2020; Zhou et al., 2020) do not need the pre-computation of SNP weights and test for inference in a two-step regression framework. Among MR based XWAS methods, summary data-based Mendelian Randomization (SMR) is among the most commonly used method (Zhu et al., 2016). It has the advantage of providing users with a heterogeneity in independent test (HEIDI) to filter out non-causal loci that may be just in linkage with causal signals.

There is widespread comorbidity among NPSUD (Plana-Ripoll et al., 2019). This is in part due to shared genetic risk factors (Lee et al., 2019), e.g., as detected by genetic correlation (rG) in cross-trait analyses (THE BRAINSTORM CONSORTIUM et al., 2018). Consequently, it is possible that there could be many shared XWAS signals among NPSUD. This supports the joint analysis of NPSUD to potentially increase detection power, especially for underpowered disorders (Turley et al., 2018; Gleason et al., 2020; Taraszka et al., 2022). Additionally, there is significant concordance of cis-eQTL and cis-mQTL effects between blood and brain (Qi et al., 2018) and more than 70% of pQTL are shared between blood and brain (Yang et al., 2021a). Moreover, the direction of effect across most tissues for shared eQTLs is the same (THE GTEX CONSORTIUM et al., 2015). Consequently, given the high comorbidities between traits and xQTL concordance between tissues, a joint analysis of the XWAS results from all traits and tissues would likely help to uncover novel signals, especially for relatively underpowered NPSUD and tissues (e.g., brain).

In this study, SMR was used to perform blood and brain XWAS of NPSUD to identify potential molecular mediators among major psychiatric disorders. To increase signal detection in underpowered disorders and tissue (brain), comorbidities among NPSUD and tissue xQTL concordances were leveraged in the joint trait/tissue concordance analysis. Subsequent analyses of XWAS signals were used to identify putative risk loci and pathways, shedding light on the etiology of NPSUD.

## 2. Materials and Methods

### 2.1 Statistical Method

We performed univariate XWAS analyses for nine NPSUD [Attention Deficit and Hyperactivity Disorder (ADHD), Autism Spectrum Disorder (ASD), Alcohol Use Disorder (AUD), Bipolar Disorder (BIP), Cannabis Use Disorder (CUD), Major Depressive Disorder (MDD), Opioid Use/Dependence Disorder (OD), Post-Traumatic Stress Disorder (PTSD) and Schizophrenia (SCZ)] (Table 1) for three paradigms (TWAS, PWAS and MWAS) and two tissues (blood and brain). For this purpose, we used SMR (v.1.03) (Zhu et al., 2016) to infer the association between the transcriptome/proteome/methylome and NPSUD. We performed SMR analysis for GWAS of NPSUD (Table 1) using external xQTL reference data sets (Table 2). To prioritize genes and perform pathway analyses, we adjusted probe (RNA/protein/CpG) SMR p-value (*P*_*SMR*_) for HEIDI test p-value (*P*_*HEIDI*_), by combining the two p-values into a single one by requiring that i) *P*_*SMR*_ is not penalized when *P*_*HEIDI*_ is above 0.01 and ii) *P*_*SMR*_ was penalized by the amount *P*_*HEIDI*_ falls below 0.01. Consequently, we adjusted *P*_*SMR*_ to 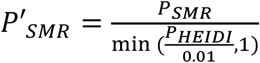. We used this approach instead of filtering by *P*_*HEIDI*_ < 0.01 because a misalignment between the GWAS cohort population and the European LD reference panel used by SMR might yield very low *P*_*HEIDI*_, e.g., the well-known C4A in our SCZ TWAS (*P*_*HEIDI*_ = 5.94*x*10^−4^) Subsequently, to extend the inference to pathways, we performed a gene set enrichment analysis for suggestive 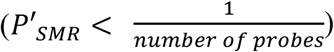 signals (Figure 1).

**Table 1.** Summary statistics of eight major PGC GWAS and MVP AUD GWAS.

| **Neuro Psychiatric and substance use disorders** | **GWAS significant markers\*/total markers** | **Study** | **Number of cases and controls / ancestry** |
| --- | --- | --- | --- |
| Attention deficit and hyperactivity disorder (ADHD) | 317 / 8,094,095 | (Demontis et al., 2019) | 19,099 - 34,194 / EUR |
| Autism spectrum disorder (ASD) | 93 / 7,822,833 | (Grove et al., 2019) | 18,381 - 27,969 / EUR |
| Alcohol use disorder (AUD) | 588 / 6,895,251 | (Kranzler et al., 2019) | 34,658 - 167,346 / EUR |
| Bipolar disorder (BIP) | 3,205 / 7,608,184 | (Mullins et al., 2021) | 41,917 - 371,549** / EUR |
| Cannabis use disorder (CUD) | 29 / 7,735,104 | (Johnson et al., 2020) | 17,193 - 357,987 / EUR |
| Major depressive disorder (MDD) | 4,625 / 7,286,335 | (Howard et al., 2019) | 411,965 - 1,285,068 / EUR |
| Opioid use/dependence disorder (OD) | 0 / 4,571,339 | (Polimanti et al., 2020) | 4,503 - 32,500 / EUR |
| Post-traumatic stress disorder (PTSD) | 3,434 / 3,875,929 | (Nievergelt et al., 2019) | 30,000 - 170,000 / EUR |
| Schizophrenia (SCZ) | 22,344 / 7,585,077 | (Trubetskoy et al., 2022) | 33,640 - 43,456 / mostly EUR |

\* Unpruned (based on LD) variants with
\*\* Case group includes bipolar or unipolar and control groups include individuals without any such diagnosis.
EUR: European

**Table 2.**
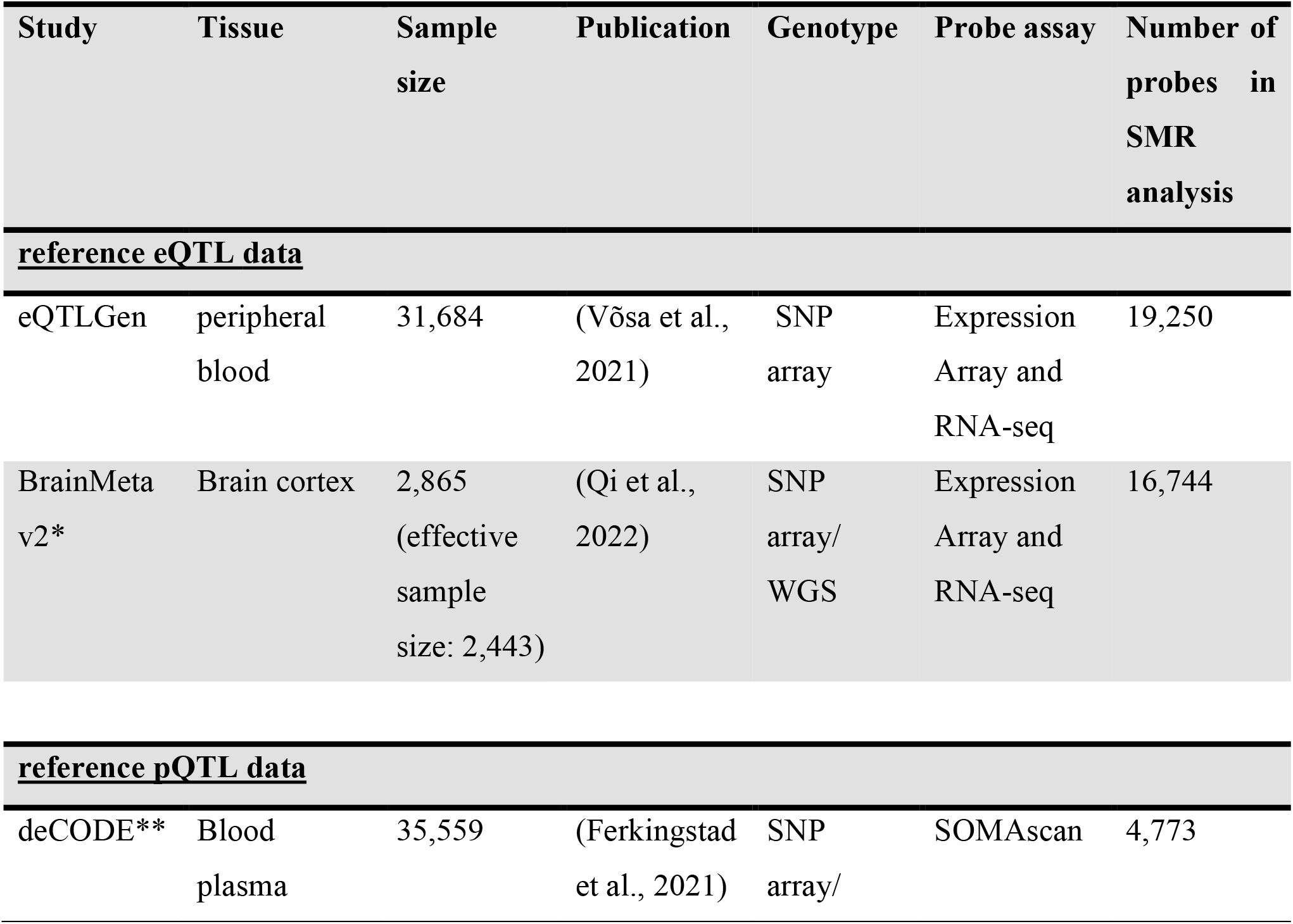
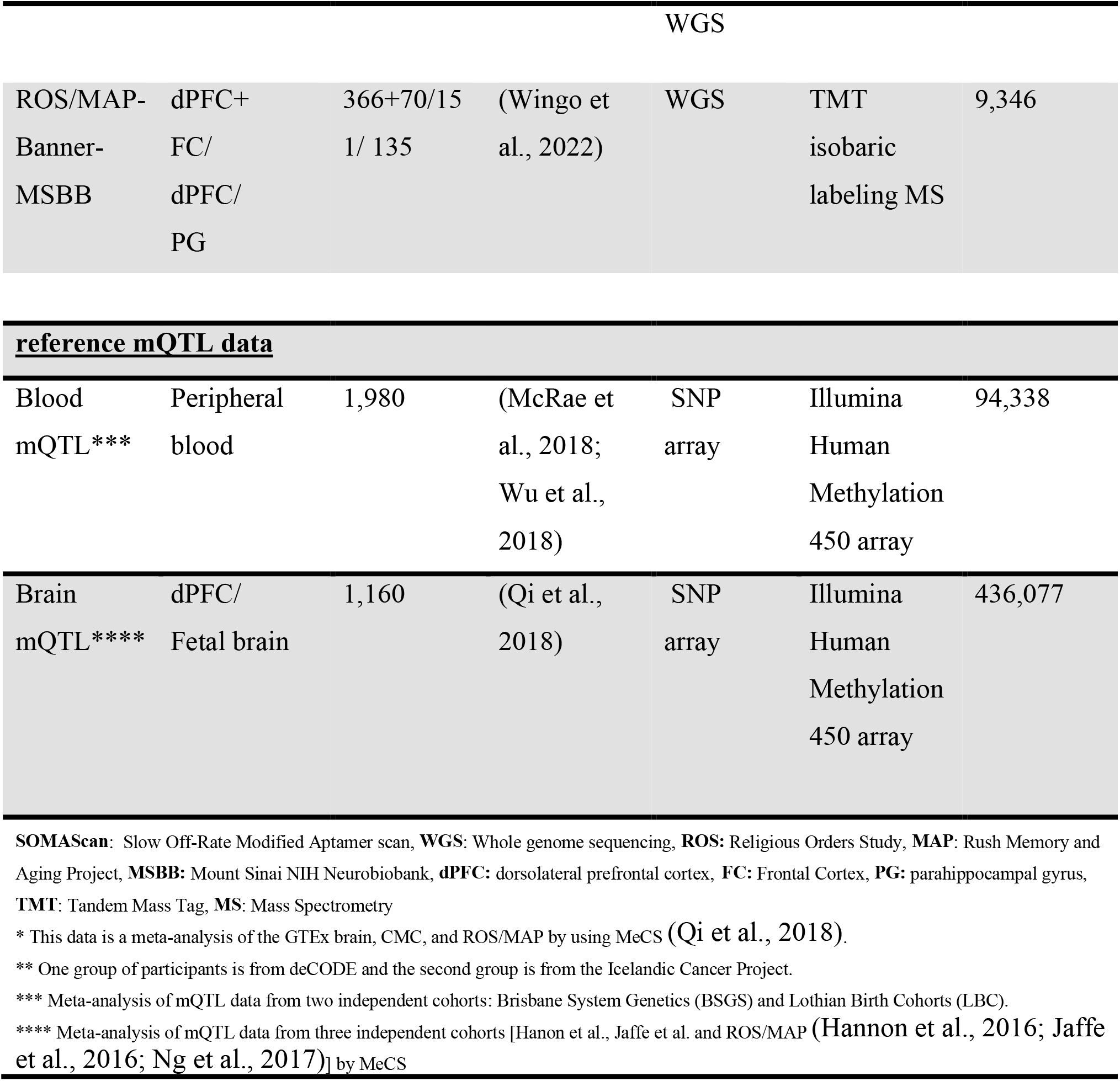
Reference xQTL molecular datasets used for XWAS studies.

**Figure 1.**
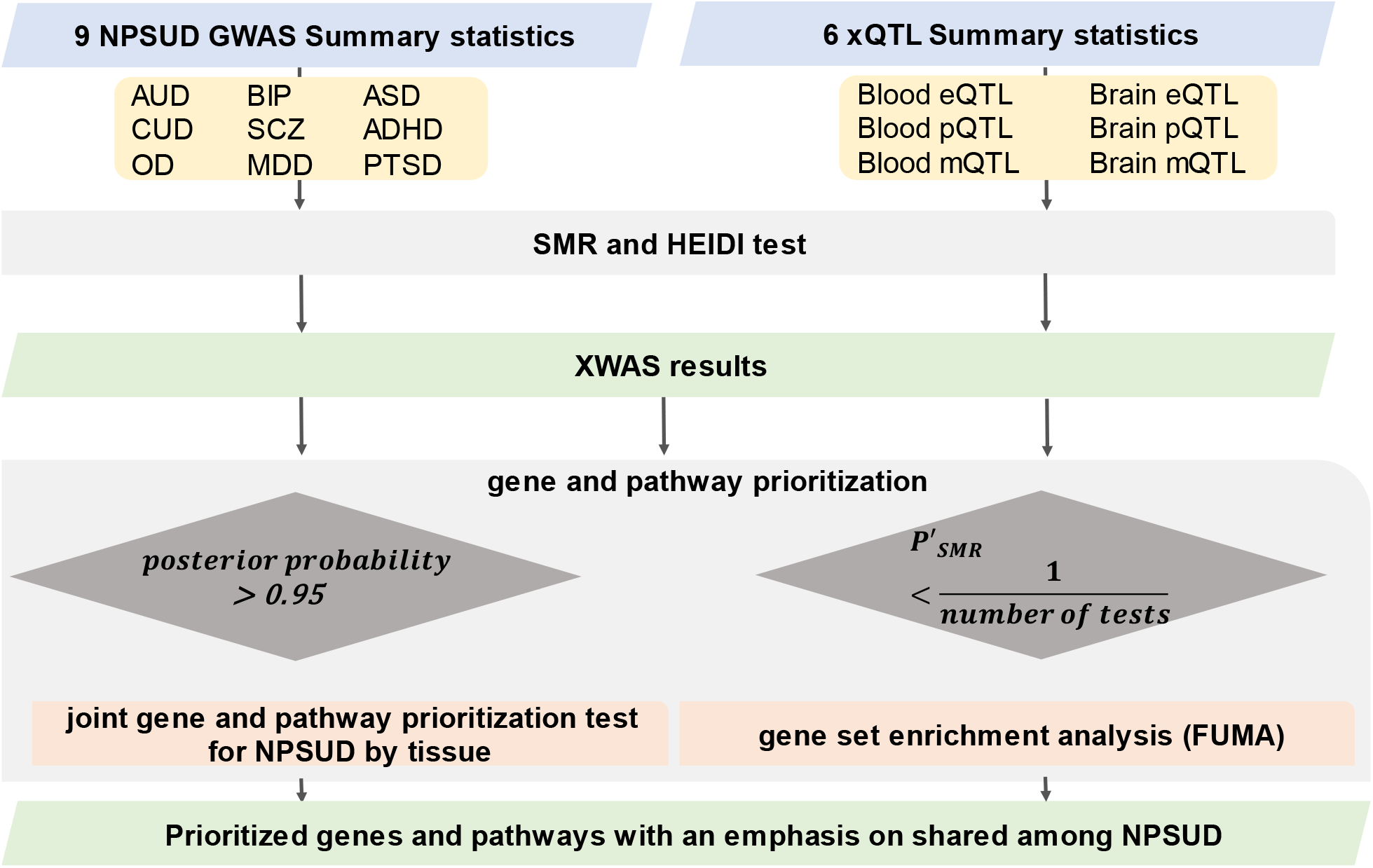
Flowchart of SMR XWAS analyses paired in both blood and brain tissues. To penalize for heterogeneity (non-causality), we employed an adjusted probe p-value [*P*′_*SMR*_ = *P*_*SMR*_/min(*P*_*HEIDI*_/0.01,1)]. For gene set enrichment analysis, we used the suggestive signals (expected to occur once per scan by chance). Primo method was used to conduct multi-trait analysis and Functional Mapping and Annotation (FUMA) was used for gene set enrichment analysis.

### 2.2 Parameters for SMR-based XWAS analyses

SMR analyses was performed for only cis-xQTLs (SNPs with p-value < 5×10^−8^ within 2 Mbp of the probe). We also used the default maximum (20) and minimum (3) number of xQTLs selected for the HEIDI test. We set the significance threshold as < 1.57×10^−3^ for xQTL p-values and the mismatch of minimum allele frequency among input files as < 15%. For HEIDI test, SNPs with LD > 0.9 and < 0.05 with top associated xQTL SNP were pruned. In case-control studies, we log-transformed the odds ratio as suggested by the SMR analysis guidelines (https://yanglab.westlake.edu.cn/software/smr/#SMR&HEIDIanalysis, accessed on August 3, 2022).

### 2.3. Neuropsychiatric and substance use disorders GWAS

All summary statistics except for AUD (Table 1) were downloaded from the Psychiatric Genomics Consortium (PGC web portal (https://www.med.unc.edu/pgc/download-results/, accessed on December 15 2023). For AUD GWAS summary statistics, we had access to the data granted through NIH from the Million Veteran Program (MVP) (dbGaP Study Accession: phs001672.v6.p1). For SMR analysis, GWAS summary statistics were processed into the SMR-ready file format. The positions for all variants and genes in the input files (LD reference panel, GWAS summary statistics, and xQTL summary statistics files) for the SMR analysis are based on the GRCh37/hg19 reference genome.

### 2.4. Molecular xQTL reference data sets

For our analyses, to get the highest signal detection, we selected the largest publicly available blood and brain xQTL datasets (Table 2). When pQTL summary statistics from reference data were not available (blood and brain pQTL) in the SMR-required input binary file format (i.e., .besd), we processed them into this .besd format. Below, we provided some of the most relevant details for these data sets. (A list of URLs for each data set is available in Supplementary Table 1, and for more details, please see the extended summary information in the Supplementary material)

#### 2.4.1. eQTL reference datasets

For TWAS, we obtained the blood eQTL data from eQTLGen (Võsa et al., 2021) and brain eQTL from BrainMeta v2 (Qi et al., 2022) (Table 2). eQTLgen consortium meta-analyzed 31,684 samples from 37 different study cohorts. Genotyping and gene expression levels were assayed mainly from whole blood (34 out of 37) and part peripheral blood mononuclear cells (3 out of 37). Most cohorts (25 out of 37) were population-based. The following eQTLGen studies included individuals of non-European ancestry (e.g., the Singapore Systems Immunology cohort -n = 115; Morocco – n = 175; Bangladeshi Vitamin E and Selenium Trial – n = 1,404). eQTLgen inferred cis-eQTL effects for 16,987 expression Genes (eGenes). BrainMeta (version 2) is a meta-analysis of brain eQTL mapping studies from seven independent cohorts (Qi et al., 2022). The study consists of 2,443 unrelated individuals of European ancestry. BrainMeta v2 detected 1,962,114 eQTL SNPs for16,744 eGenes.

#### 2.4.2. pQTL reference datasets

For PWAS, we used the blood pQTL data from deCODE (Ferkingstad et al., 2021) and brain pQTL from Wingo et al. (Wingo et al., 2022) (Table.2). The deCODE proteome study consisted of 35,559 individuals from Iceland. Blood plasma samples were assayed for 4,907 probes [Slow Off-rate Modified Aptamer Scan (SOMAScan) (Gold et al., 2010, 2012) assay version 4 aptamers], which correspond to 4,719 unique proteins.

The brain pQTL study sampled three regions of the brain: prefrontal cortex, dorsolateral prefrontal cortex and parahippocampal gyrus (Wingo et al., 2022) in 722 samples. It used isobaric tandem mass tag method to assay proteins and 9,363 of them met the quality control criteria. While the sample size of brain pQTL reference data is relatively small, this was the largest publicly available such study at the time of completion for the analyses.

#### 2.4.3. mQTL reference datasets

For MWAS analyses, we used the blood (McRae et al., 2018; Wu et al., 2018) and brain mQTL data sets (Qi et al., 2018) (Table.2), which are publicly available for download from SMR web portal (https://yanglab.westlake.edu.cn/software/smr/#DataResource, accessed on January 28 2023). The mQTL data for the brain is a meta-analysis of the mQTL mapping results from three major studies (Hannon et al., 2016; Jaffe et al., 2016; Ng et al., 2017). The methylation assay used in these studies was the Illumina Infinium Human Methylation 450K array. We used the annotation file provided by the manufacturer to map the CpG probe ids (with “cg” prefix) to the HUGO gene nomenclature committee (HGNC) gene symbol.

### 2.5. Gene set enrichment analysis

Our aim is to uncover pathways that are associated with NPSUD. For this purpose, we tested for pathway enrichment in XWAS signals. We performed two separate Functional Mapping and Annotation (FUMA) (v1.41) (Watanabe et al., 2017) pathway analyses using genes with suggestive signals from: i) TWAS and PWAS combined and ii) MWAS only. We included genes with suggestive adjusted p-values () in query gene lists for FUMA analyses assumed a possible universe of 54,619 coding and non-coding genes (protein coding, long non-coding RNA, non-coding RNA and processed transcripts). Due to the complexity of the MHC region, we chose the option for excluding genes from this region in FUMA analyses. To adjust for multiple testing, we assessed pathway significance using the False Discovery Rate (FDR) procedure (q-value < 0.05) (Benjamini and Hochberg, 1995).

### 2.6 Joint NPSUD concordance analysis of TWAS and MWAS gene signals

To increase the statistical power for the prioritization of genes in underpowered NPSUD and tissues (such as brain), we used a multi-trait and multi-tissue approach. Therefore, we conducted a joint trait concordance analysis using Primo (R package for Integrative Multi-Omics association analysis) (Gleason et al., 2020) within the more powerful XWAS paradigms (TWAS and MWAS). We did not jointly analyze PWAS because brain results were too sparse. We used Primo because it was designed to jointly analyze summary statistics from multiple studies while adjusting for the correlation between datasets (e.g., due to sample overlapping). Gene-level adjusted p-values from SMR analyses were used as input for the joint trait and tissue concordance analyses. If a gene had multiple p-values then the Cauchy method (Liu et al., 2019) was used to combine these p-values to have one p-value for the gene. Because Primo requires the estimated proportion of statistics (alt_props) coming from the alternative distribution, we exhaustively tested different values for this parameter. We also estimated it directly from the data using a mixture of two distributions. This parameter was critical because more significant results were identified when larger values of alt_probs were used. We finally decided to set alt_probs=10^−3^ which was also suggested in the Primo paper (Gleason et al., 2020). For prioritization purposes, we considered genes with posterior probabilities (PP) > 0.95 as significant.

Besides increasing signal detection in the brain, the joint analysis might open the avenue for further investigations. For instance, blood and brain concordant signals might be further studied to be used as proxies for the brain pathology of NPSUD. It is possible that such blood markers might also have an impact on the diagnosis/prognosis of NPSUD, i.e., concordant XWAS signals might have translational implications.

## Results

In this section, we provide a selection of the common and shared XWAS results. Due to the strong signals in MHC region making the visualization of other findings difficult, we omitted this region from the plots of results coming from univariate TWAS and PWAS analyses (Figure 1 and 2). Details on MHC signals for these two paradigms were provided in Supplementary Figures 1 and All univariate XWAS results collated by paradigm are available in Supplementary data files.

**Figure 2.**
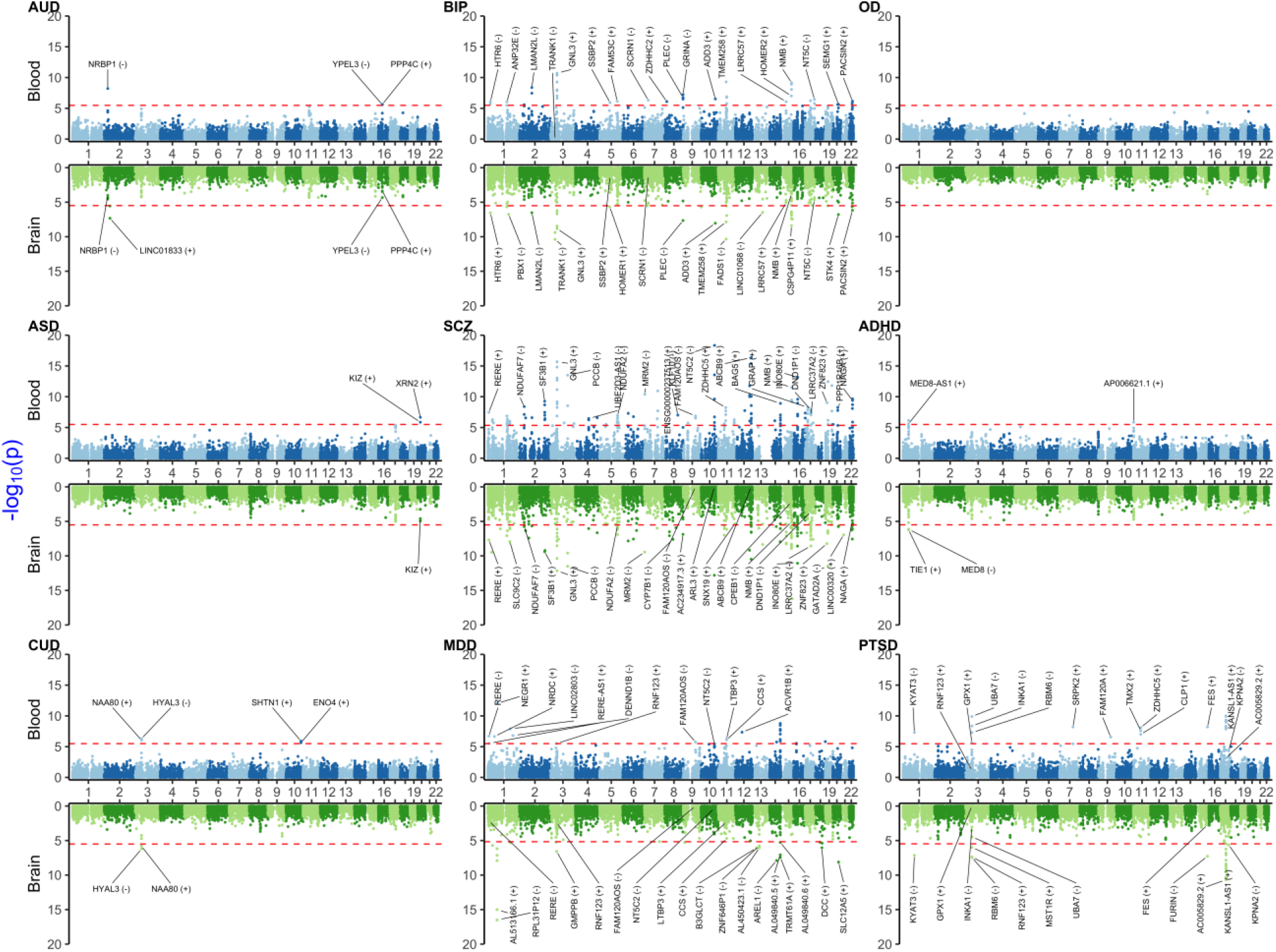
Miami plot [Manhattan blood (above)-brain (below) bi-plot] of TWAS adjusted p-values (*P*′_*SMR*_) for investigated neuropsychiatric and substance use disorders. The upper part of the plot is for the blood and the lower part is for the brain. The red horizontal line denotes Bonferroni significance threshold. For visualization, we labeled the signals by their affiliated HUGO gene name and the direction of the SMR effect estimate on the trait shown in parentheses. **AUD:** Alcohol Use Disorder **BIP:** Bipolar Disorder **OD:** Opioid Dependence/Use Disorder **ASD:** Autism Spectrum Disorder **SCZ:** Schizophrenia **ADHD:** Attention Deficit and Hyperactivity Disorder **CUD:** Cannabis Use Disorder **MDD:** Major Depression **PTSD:** Post-traumatic stress disorder.

### TWAS results

Blood and brain TWAS for BIP, SCZ and MDD yielded the highest number of significant TWAS signals (Figure 2 and Supplementary Figure 1). These disorders share many common signals, especially in the Major Histocompatibility Complex (MHC) region on chromosome 6 (25-35 Mbps), e.g., *BTN3A2* and *C4A*, which were concordant (i.e., significant and with the same sign for effect size) between blood and brain. Other shared signals between three disorders were *ATF6B, C4A, FLOT1, IER3, LINC00243, TRIM10, TUBB, TNXA, ZNF602P* and *ZSCAN12P1*, in blood and, *OR2B8P, ZKSCAN8P1* and *ZSCAN16-AS1* in brain.

Among substance use disorders (SUD), AUD showed significant signals in blood (*NRBP1, PPP4C* and *YPEL3*) and brain (*LINC01833*). For CUD, *HYAL3* and *NAA80* on chromosome 3 were significant signals and with concordant direction of effect between blood and brain. CUD also had significant blood-only signals on chromosome 10 in *ENO4* and *SHTN1*. However, robust signals were not detected for OD.

For ADHD, there were three significant signals from blood (*AL139289.1, AP006621.1* and *MED8-AS1*) and two from brain (*MED8* and *TIE1*). For ASD, there were two significant signals on chromosome 22 (*KIZ* and *XRN2*), with *KIZ* also being suggestive in brain. We observed signals for PTSD on chromosome 17, some of which had concordant direction of effect between blood and brain (e.g., *AC005829.23* and *KANSL1-AS1*). PTSD results also yielded a blood-brain concordant signal for *KYAT3*.

### PWAS results

When compared to TWAS, there was a lower number of PWAS significant signals (Figure 3 and Supplementary Figure 2). This is expected because PWAS had a lower number of probes tested and lower sample for reference panels, especially for brain tissue. For instance, we did not identify any significant blood or brain PWAS signals for OD, ASD or CUD. Similar to TWAS - BIP, SCZ and MDD yielded common signals in MHC regions for blood (*BTN3A3, BTN3A1* and *MICB* - see also Supplementary Figure 2 for more details). *NEK4* was a brain only signal shared between SCZ and BIP.

**Figure 3.**
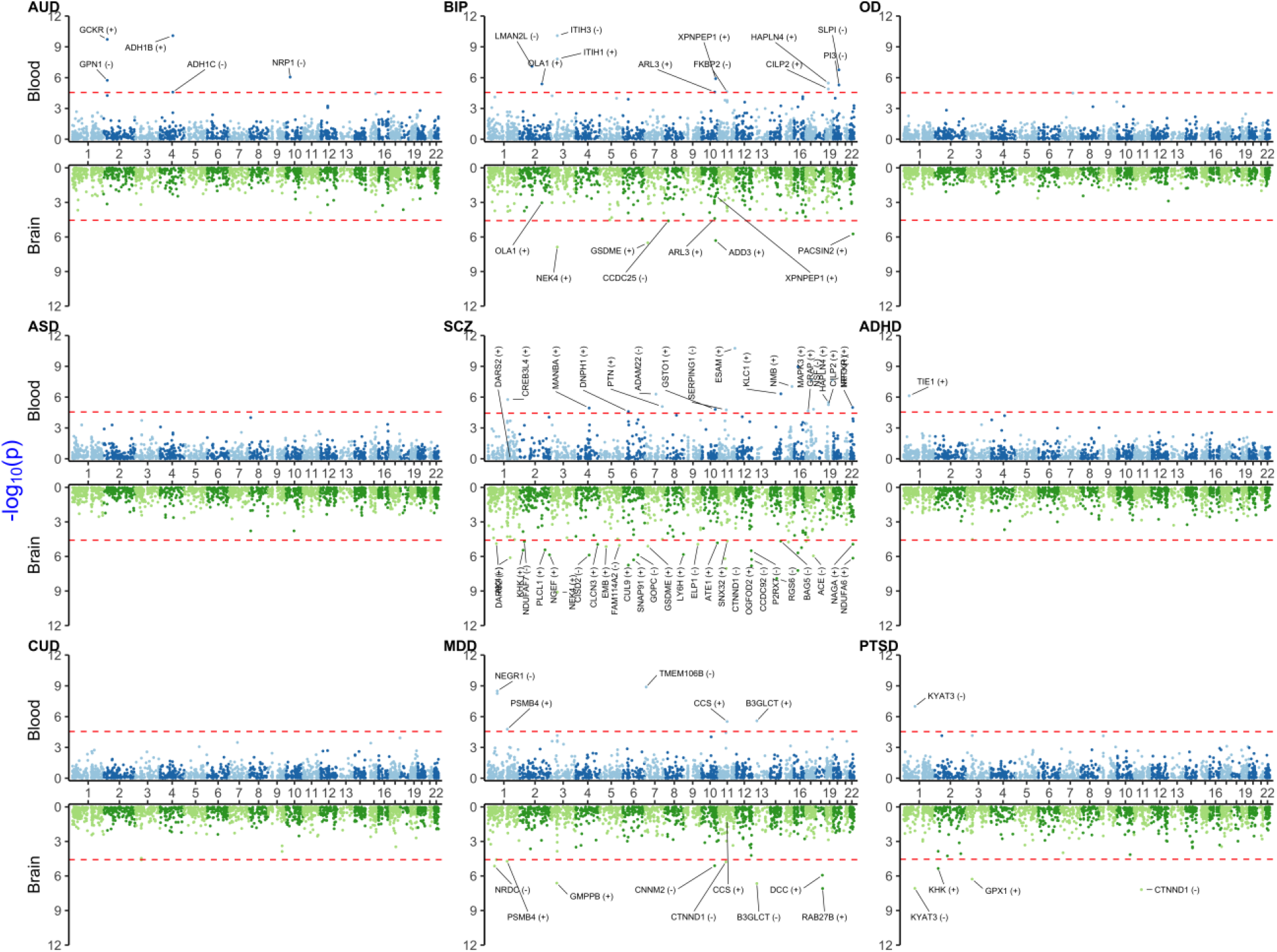
Miami plot [Manhattan blood(above)-brain (below) bi-plot] of PWAS adjusted p-values (*P*′_*SMR*_) for investigated neuropsychiatric and substance use disorders. For details and background see Figure 2 legend.

For PTSD, significant brain signals were in *KYAT3* (detected also in TWAS), *KHK, GPX1, MICB* and *CTNND1* (that is also common signal between SCZ and MDD). Among these, only *KYAT3* was blood-brain concordant for the direction of effect. Some notable disease specific signals were for *ADH1C* and *ADH1B* in AUD blood and *TIE1* in ADHD blood PWAS (which was also significant in ADHD brain TWAS).

### MWAS results

Notably, biologically significant signals were detected, e.g., *ADH1C* for AUD in blood. Similar to TWAS and PWAS, we found that BIP, SCZ and MDD had more significant signals than the remaining disorders (Figure 4 and Supplementary Files). Again, the largest blood-brain concordant signals that were common between BIP, SCZ and MDD were in the MHC region, such as, *ZSCAN12L1, BTN3A2, H2AC13, ZNF389* for brain and *VARS2, TUBB, TRIM27, TRIM31, DPCR1, TRIM15, DDR1, GTF2H4, TRIM40, H2AC13, TRIM26, PBX2, HIST1H4D, SFTA2, BTN3A2, HCG9, HLA-B* and *MSH5* for blood.

**Figure 4.**
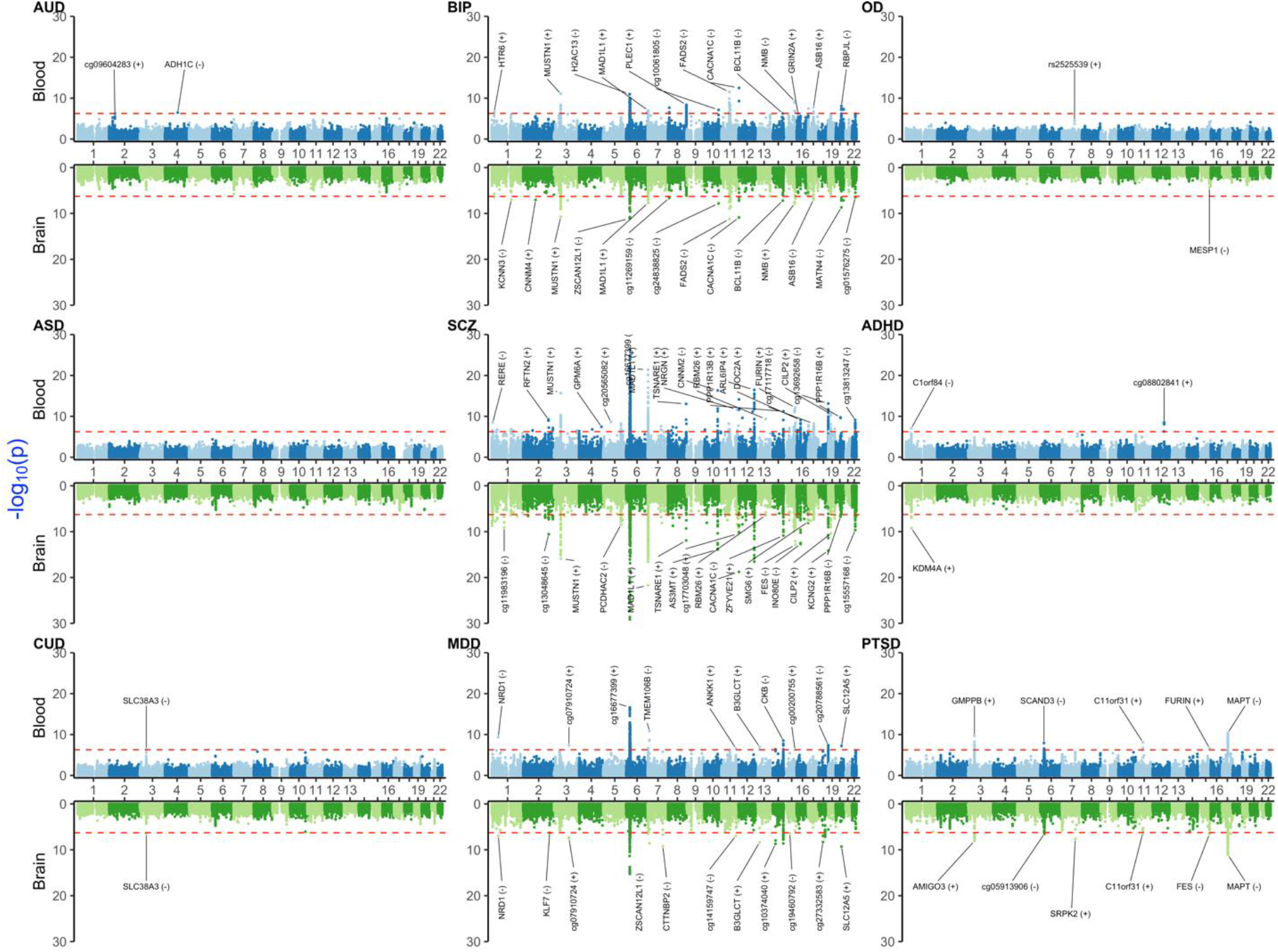
Miami plot [Manhattan blood (above)-brain (below) bi-plot] of MWAS adjusted p-values (*P*′_*SMR*_) for investigated NPSUD. For details and background, see Figure 2 legend.

As in PWAS, there were no significant signals for OD or ASD. For CUD, *SLC38A3* was a blood-brain concordant signal. ADHD yielded two significant signals on chromosome 1 (*C1orf84* in blood and *KDM4A* in brain). For PTSD, there were significant signals for *GMPPB, SCAND3, C11orf31, FURIN* and *MAPT* in blood. Among these, *MAPT* and *C11orf31* were also concordant between blood and brain. Other leading PTSD brain signals were *AMIGO3*, cg05913906 and *FES*.

### Gene set enrichment analysis results

In this section, we highlighted some of the more interesting signals from FUMA gene set enrichment. More detailed results can be found in Supplementary Figures 3-33 and in Supplementary Tables 2 and 3. Consistent with most other XWAS results, there were no significant findings for OD and CUD. As expected, i) SCZ yielded the highest number of signals (due to its larger sample size in GWAS) and ii) alcohol metabolism pathways showed significant enrichments for AUD. For BIP, the combined TWAS and PWAS prioritized genes were significantly enriched in non-genomic actions of the 1,25 dihydroxyvitamin D3 gene set (*PLCB3, PRKCB, PRKCA* and *CD40*) (q-value = 3.81×10^−2^) (Supplementary Figure 10). Another BIP signal was GWAS catalog gene enrichment for plasma omega-3 polyunsaturated fatty acid levels (alpha-linolenic acid) (*MYRF, TMEM258* and *FADS1*) (q-value = 1.00 ×10^−3^) (Supplementary Figure 12). For the same disorder, GO_HYALURONAN_METABOLIC_PROCESS (*ITIH1, ITIH3* and *ITIH4*) (q-value = 5.67×10^−3^) was the most significant Gene Ontology (GO) terms in the Biological Process category (Supplementary Figure 13). For more details, refer to Supplementary Tables 2 and 3. Peptidase related GO terms were shared signals between BIP, MDD and SCZ (Supplementary Figures 34-35). Also, neuron related pathways (GO_SYNAPSE_PART, GO_PRESYNAPSE and GO_POST_SYNANSE) were significant enrichment for MDD (Supplementary Figures 34 and 35).

In the MWAS only analysis, again SCZ yielded the most signals. Metabolism of alpha-linolenic acid (omega-3) was one of the significant gene sets for BIP blood MWAS (*FADS2, FADS1* and *MIR1908*) (q-value = 3.24×10^−4^) (Supplementary Figure 33). BIP and SCZ shared a signal for cation ion transport related gene sets (GO_CATION_TRANSPORT and GO_DIVALENT_INORGANIC_CATION_TRANSPORT) (Supplementary Figure 37). See Supplementary Tables 4 and 5 for details.

### Joint analysis for both i) NPSUD and ii) blood and brain

TWAS/MWAS results were jointly analyzed for seven NPSUD (Figure 5), excluding the underpowered OD and CUD due to poor distributions of XWAS p-values vs Primo PPs.

**Figure 5.**
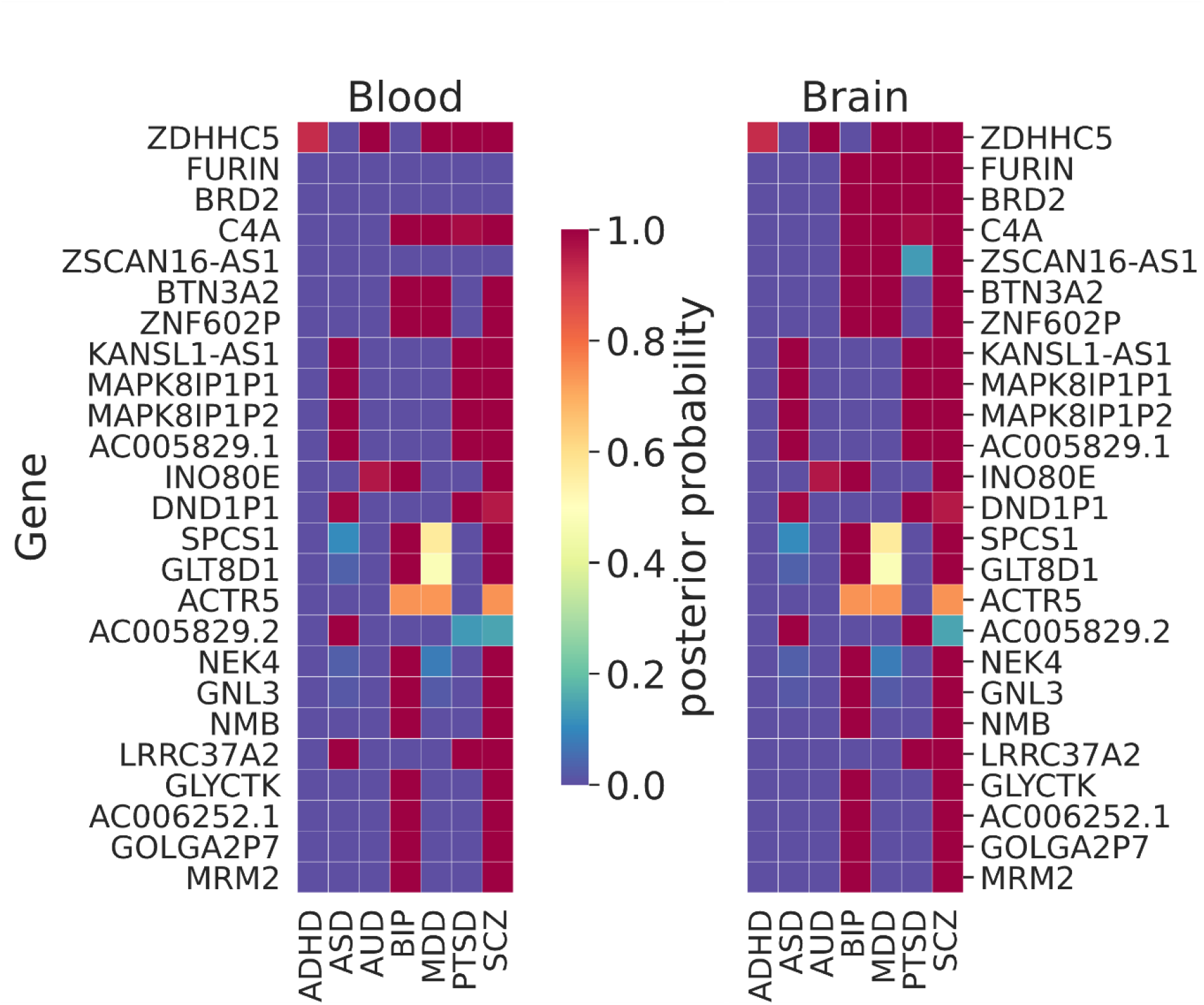
Results of the joint trait concordance analysis for TWAS. The top twenty-five genes are shown as ranked by the sum of the posterior probabilities (PPs) within brain tissue for disorders: Attention deficit and hyperactivity disorder (ADHD), Autism spectrum disorder (ASD), Alcohol use disorder (AUD), Bi-polar disorder (BIP), Major depressive disorder (MDD), Post-traumatic stress disorder (PTSD) and Schizophrenia (SCZ).

We observed gene signals (PP>0.95) that are shared between many NPSUD and between blood and brain (Figure 5). *ZDHHC5* was the most shared signal between blood and brain and five NPSUD (ADHD, AUD, MDD, PTSD and SCZ). There is a cluster of genes that is also shared only between ASD, PTSD and SCZ, e.g., *LRRC37A2, KANSL1-AS1, MAPK8IP1P1, MAPK8IP1P2* and *AC005829.1*. BIP and SCZ also share a number of signals (*NEK4, GNL3, NMB, GLYCTK, AC006252.1* and *GOLGA2P7)*.

However, there were also disease specific genes, e.g., i) *AP006621.3* and *PIDD1* for ADHD, ii) *ADD3, LMAN2L* and *PLEC* for BIP, iii) *KYAT3* and *PLEKHM1* for PTSD, iv) *PCCB* and *GATAD2A* for SCZ and v) *LINC02803* for MDD. (Detailed in Supplementary Table 6)

While often there were very similar pattern of shared TWAS signals between blood and brain, there were also brain specific signals. For instance, *BRD2, FURIN* and *ZSCAN16-AS1* were such brain only signals that are shared among many disorders. [Note that FURIN was successfully tested, via CRISPR/Cas9 experiment on isogenic human induced pluripotent cell, for the allelic effect on its gene expression of the SNP with the largest SCZ signal in the region (Schrode et al., 2019).] There were also both disease and brain specific signals, e.g., *FTCDNL1* for SCZ observed only in brain. For more disease/tissue specific signals, please see also Supplementary Table 6.

For MWAS, we often observed the same pattern of shared signals between blood and brain. Among the largest signals (ranked by the sum of PP in brain), *C11orf31, MED19* and *FURIN* were shared among ADHD, AUD, BIP, MDD, PTSD and SCZ (Figure 6). *GATAD2A* stood out as shared brain specific signal among ADHD, ASD, BIP, PTSD and SCZ. *RERE* was shared among AUD, MDD, PTSD and SCZ, which was one of the eGenes for a cis-eQTL associated with SCZ that showed allele specific effect via the chromatin interaction (Zhang et al., 2020). For more details about results, see Supplementary Table 7.

**Figure 6.**
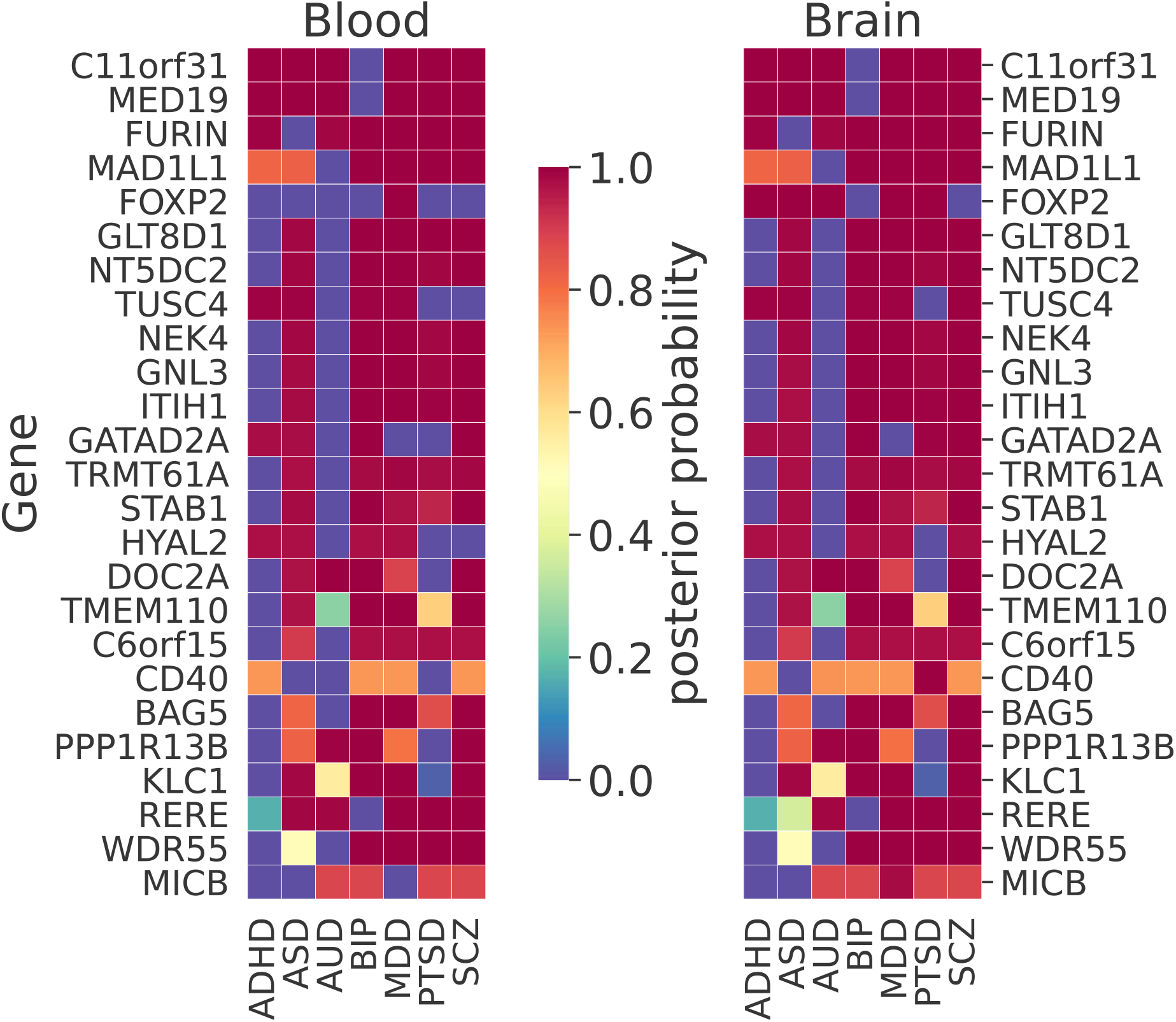
Results of joint trait concordance analysis for MWAS. The top twenty-five genes (excluding probes not mapped to a gene name as per Illumina annotation) are shown and ranked by sum of brain PPs for all disorders.

## Discussion

To provide biological context to GWAS findings, we performed XWAS analyses using the SMR was performed. These analyses uncovered putative risk genes by inferring the association between the transcriptome/proteome/methylome and NPSUD. We subsequently identified molecular pathways associated with NPSUD via gene set enrichment analyses of genes that yielded XWAS suggestive signals. To improve signal detection power for underpowered traits and brain tissue, we also performed a joint concordance analysis of all traits and tissues within the two adequately powered XWAS paradigms (TWAS and MWAS) were performed. The results of this work suggest possible components of the treatment regimen for certain NPSUD, e.g., the possible implication of vitamins (B6 and D) and omega-3 pathways for some of these disorders.

Our analyses replicated biologically relevant and previous significant findings. Among the biologically relevant ones, we note that the common signal in AUD between blood PWAS and MWAS was *ADH1C*, which codes for alcohol dehydrogenase enzyme that metabolizes the alcohol. It was also implicated as a significant loci in various GWAS of alcohol related phenotypes (Gelernter et al., 2014; Clarke et al., 2017; Kranzler et al., 2019). We also replicated findings from Dall’Aglio et al. (Dall’Aglio et al., 2021), in which *NEGR1* was found to be a signal for MDD as it was the second most significant gene in our MDD blood TWAS. Similar to our TWAS results, *BTN3A2* and *RPL31P12* were significant findings in a previous MDD brain TWAS from Yang et al. (Yang et al., 2021b).

In our joint TWAS concordance analyses, *ZDHHC5* was the shared signal between all NPSUD except for ASD and BIP (Figure 5). This gene was previously found to be a shared blood TWAS signal between MDD and SCZ (Reay and Cairns, 2020). In the same paper, some of our other XWAS signals shared between BIP and SCZ (e.g., *NEK4, GNL3* and *NMB*) were also reported as shared TWAS signals in blood. Another shared signal between SCZ, BIP and AUD was *INO80E* which was previously indicated as one of the top ten shared signals between SCZ and AUD (Johnson et al., 2021). A common blood-brain MWAS signal for AUD, MDD, PTSD and SCZ from the Primo joint analyses was *RERE*. It was one of the eGenes for a cis-eQTL showing allele specific effect via the chromatin interaction (Zhang et al., 2020). The same gene was also implicated as a significant gene in SMR analysis in the recent PGC SCZ GWAS (Trubetskoy et al., 2022).

We compared our joint trait concordance analysis findings for SCZ (brain TWAS+MWAS or brain TWAS only) with PGC3 SCZ GWAS findings. There were common genes identified as significant (PP>0.95) in our brain tissue results (TWAS+MWAS) and those found in the list of significant replication and discovery loci (see Supplementary Table 8) in PGC3 SCZ GWAS, such as, *BTN3A2, FURIN, GATAD2A, GNL3, INO80E, KANSL1-AS1* and *NEK4*. However, we found significant signals for *AC005829.1, BRD2, C11orf31, C6orf15, DND1P1, MAPK8IP1P1* and *ZNF602P*, which were not found as significant in the above PGC3 SCZ gene list. There are also common genes between our brain tissue TWAS results (from the joint analysis) and the list of genes in PGC SCZ that were prioritized based on the SMR analysis only. Those genes are *INO80E, GATAD2A, PCCB* and *FURIN*. However, our brain results include significant findings that were not identified in PGC3 SCZ, such as *BRD2, GNL3, KANSL1-AS1, NEK4* and *ZDHHC5*. (For details see Supplementary Table 8)

The MHC region is a well-known region associated with some of the NPSUD, e.g., SCZ (Corvin and Morris, 2014). Our joint XWAS analyses strongly support this assertion for SCZ, BIP, PTSD and MDD. For instance, *BTN3A2* was the leading TWAS signal for BIP, MDD and SCZ. Also, *C4A*, thought to be implicated in synaptic pruning (Sekar et al., 2016), was a shared brain TWAS signal for BIP, MDD, PTSD and SCZ (Figure 5). Similarly, *MICB* was shared between AUD, BIP, MDD, PTSD and SCZ in brain MWAS (Figure 6). Its possible involvement in NPSUD was also supported by empirical transcriptomic evidence. Research showed that *MICB* is part of a molecular network interacting with the differentially expressed genes in Brodmann’s area 9 region of individuals with MDD (Scarr et al., 2019).

Beside many common signals between NPSUD, there are also some that are disease specific. For PTSD, we observed specific signals for TWAS/PWAS in a cluster of genes on chromosome 17 and *KYAT3* on chromosome 1. *KYAT3* was the strongest signal for PTSD brain PWAS. It was reported as a TWAS signal for reexperiencing a PTSD symptom cluster (Pathak et al., 2022). A GWAS on social anxiety also found SNP that is downstream of KYAT3 to be significant (Stein et al., 2017).

AUD specific TWAS signals were on chromosome 2 for *NRBP1* and *SNX17*. A meta-analysis of Alcohol Use Disorders Identification Test (AUDIT) showed that index SNP for *GCKR* overlap also with the *SNX17* (Sanchez-Roige et al., 2019). This same gene was also found significant (p-value = 1.18×10^−6^; brain caudate basal ganglia) in TWAS of substance use disorder (Hatoum et al., 2022). For ADHD, *TIE1* (chromosome 1) is a TWAS/PWAS signal that was not found in other NPSUD. This gene is coding for tyrosine kinase and it was found as significant in PTSD TWAS (Liao et al., 2019) and ADHD TWAS (Chen et al., 2022). For CUD, we also identified concordant blood-brain TWAS signals on chromosome 3, e.g., *HYAL3* and *NAA80*. In another TWAS analysis using the same CUD GWAS (Table 1), they also found *HYAL3* was found to be significant (Johnson et al., 2020). Previously, expression of *NAA80* in brain (anterior cingulate cortex) was associated with the genome-wide significant variant rs2777888 in meta-analyzed European ancestry PTSD GWAS (Gelernter et al., 2019).

Since blood and brain cis-xQTL were known to overlap, this was used to improve the discovery power for brain XWAS analyses through the joint analysis of both tissues. The joint analyses found many candidate risk genes that are concordant in direction of effect for both tissues. The blood XWAS of these concordant genes might be useful for future development of NPSUD multivariate blood biomarkers that might be used for diagnosis, prognosis and possible treatment of these disorders.

Supplements such as vitamins and omega-3 have been tested (with varying success rates) as treatment for NPSUD (Firth et al., 2019). Previous investigations in blood indicated that the deficiency (Anglin et al., 2013) and supplementation (Sarris et al., 2016) of vitamin D might increase/decrease the risk for MDD. Other studies did not find any effect of vitamin D supplementation on MDD (Marsh et al., 2017) and depression in older adults (Okereke et al., 2020). A most recent GWAS of polyunsaturated and monounsaturated fatty acids showed a significant gene set enrichment of GWAS catalog genes in Bipolar I and II (Francis et al., 2022). Omega-3 was also found to likely lower MDD risk (Mocking et al., 2016). It is not clear if these non-genetic/non-MR studies did eliminate most confounders. However, most confounders are eliminated by using mendelian randomization methods, such as SMR that was used in this work.

While we did not uncover any vitamin associated pathway or gene signal for MDD, our analyses indicated link between such supplements and other NPSUD. For instance, BIP TWAS and PWAS prioritized genes showed significant gene set enrichment for the vitamin D3 pathway (Supplementary Figure 10). For BIP, we also found significant gene set enrichment in omega-3 associated gene sets in combined T/PWAS (Supplementary Figure 11) and MWAS only (Supplementary Figure 36) FUMA analyses. *KYAT3* signal found in PTSD T/PWAS suggests a possible etiological role of vitamin B6 in this disease. Based on these findings, more clinical research evidence is needed to test these molecules as a secondary component of the treatment regimen: i) vitamin D and omega-3 modest supplementation for BIP and ii) vitamin B6 (or B complex) for PTSD.

The importance of this study is five-fold. First, this is the most powerful XWAS study of NPSUD because we integrated the largest available xQTL reference data and NPSUD GWAS. Second, we extended these most powerful gene-level XWAS inferences to pathway level, which suggested some novel avenues for treatment. Third, we further increased the detection power for underpowered trait and tissues via multi-trait multi-tissue joint analyses. Fourth, the joint analyses uncovered blood-brain concordant XWAS signal that, in the future, might form the basis for the development of (multivariate) blood biomarkers for diagnosis/prognosis. Fifth, these joint analyses are the first formal attempt to uncover common signals for multiple disorders and those specific to a single one.

## Limitations of the study

1. Although we applied the HEIDI test, it is not likely that SMR completely eliminates the horizontal pleiotropy. For instance, a SNP might be xQTL for multiple genes, which violates the assumption that the SNP effect on the trait is mediated only through the tested gene. However, we believe that gene set/pathway inferences are likely to mitigate the confounding effect of this phenomenon from gene-level analysis.

2. While Primo can adjust for the correlations between multiple different studies, it does not correct for the correlation between genes (e.g., which can happen due to the linkage disequilibrium of variants). Thus, future studies need to validate the identified genes.

3. Reference brain pQTL has lower sample sizes than the blood pQTL data. This resulted in fewer significant signals for brain PWAS.

4. Some reference xQTL data (e.g., brain proteome) are enriched in individuals with certain neurological disorders.

5. We used 1000 Genome phase 3 as a reference LD panel that might not be an exact match for the LD patterns from the GWAS cohort. By adjusting *P*_*SMR*_, we eliminated the inflation of *P*_*HEIDI*_, signals due to any cohort-panel LD mismatch.

6. Due to extended and irregular LD patterns, findings in certain regions (e.g., MHC) should be interpreted with care.

7. Since we only included xQTL reference data obtained from bulk tissue, any cell type specific information was not presented in our findings.

## Data availability statement

All publicly available data used in this study were listed in Supplementary Table 1. All the data generated in this study will be open to public when the manuscript is published.

## Data Availability

All data produced in the present study are available upon reasonable request to the authors

## Authors’ Contributions

HG completed the XWAS analysis and data visualization. THN conducted the joint trait analysis. CC contributed GWAS data. VIV, BPR and SAB supervised the study design. All authors contributed to the preparation of the manuscript.

## References

Aberg, K. A., Dean, B., Shabalin, A. A., Chan, R. F., Han, L. K. M., Zhao, M., et al. (2020). Methylome-wide association findings for major depressive disorder overlap in blood and brain and replicate in independent brain samples. Mol Psychiatry 25, 1344–1354. doi: 10.1038/s41380-018-0247-6.

Anglin, R. E. S., Samaan, Z., Walter, S. D., and McDonald, S. D. (2013). Vitamin D deficiency and depression in adults: systematic review and meta-analysis. The British Journal of Psychiatry 202, 100–107. doi: 10.1192/bjp.bp.111.106666.

Aygün, N., Elwell, A. L., Liang, D., Lafferty, M. J., Cheek, K. E., Courtney, K. P., et al. (2021). Brain-trait-associated variants impact cell-type-specific gene regulation during neurogenesis. The American Journal of Human Genetics 108, 1647–1668. doi: 10.1016/j.ajhg.2021.07.011.

Bae, Y. E., Wu, L., and Wu, C. (2021). InTACT: An adaptive and powerful framework for joint-tissue transcriptome-wide association studies. Genet Epidemiol. doi: 10.1002/gepi.22425.

Barbeira, A. N., Dickinson, S. P., Bonazzola, R., Zheng, J., Wheeler, H. E., Torres, J. M., et al. (2018). Exploring the phenotypic consequences of tissue specific gene expression variation inferred from GWAS summary statistics. Nat Commun 9, 1825. doi: 10.1038/s41467-018-03621-1.

Barbeira, A. N., Pividori, M., Zheng, J., Wheeler, H. E., Nicolae, D. L., and Im, H. K. (2019). Integrating predicted transcriptome from multiple tissues improves association detection. PLOS Genetics 15, e1007889. doi: 10.1371/journal.pgen.1007889.

Benjamini, Y., and Hochberg, Y. (1995). Controlling the False Discovery Rate: A Practical and Powerful Approach to Multiple Testing. Journal of the Royal Statistical Society: Series B (Methodological) 57, 289–300. doi: 10.1111/j.2517-6161.1995.tb02031.x.

Brennand, K., Simone, A., Tran, N., and Gage, F. (2012). Modeling psychiatric disorders at the cellular and network levels. Mol Psychiatry 17, 1239–1253. doi: 10.1038/mp.2012.20.

Bryois, J., Calini, D., Macnair, W., Foo, L., Urich, E., Ortmann, W., et al. (2022). Cell-type-specific cis-eQTLs in eight human brain cell types identify novel risk genes for psychiatric and neurological disorders. Nat Neurosci 25, 1104–1112. doi: 10.1038/s41593-022-01128-z.

Buenrostro, J. D., Wu, B., Litzenburger, U. M., Ruff, D., Gonzales, M. L., Snyder, M. P., et al. (2015). Single-cell chromatin accessibility reveals principles of regulatory variation. Nature 523, 486–490. doi: 10.1038/nature14590.

Chen, X., Yao, T., Cai, J., Zhang, Q., Li, S., Li, H., et al. (2022). A novel cis-regulatory variant modulating TIE1 expression associated with attention deficit hyperactivity disorder in Han Chinese children. Journal of Affective Disorders 300, 179–188. doi: 10.1016/j.jad.2021.12.066.

Clarke, T.-K., Adams, M. J., Davies, G., Howard, D. M., Hall, L. S., Padmanabhan, S., et al. (2017). Genome-wide association study of alcohol consumption and genetic overlap with other health-related traits in UK Biobank (N=112 117). Mol Psychiatry 22, 1376–1384. doi: 10.1038/mp.2017.153.

Consortium, T. Gte. (2020). The GTEx Consortium atlas of genetic regulatory effects across human tissues. Science. Available at: http://www.science.org/doi/abs/10.1126/science.aaz1776 [Accessed September 6, 2021].

Corvin, A., and Morris, D. W. (2014). Genome-wide Association Studies: Findings at the Major Histocompatibility Complex Locus in Psychosis. Biological Psychiatry 75, 276–283. doi: 10.1016/j.biopsych.2013.09.018.

Dall’Aglio, L., Lewis, C. M., and Pain, O. (2021). Delineating the Genetic Component of Gene Expression in Major Depression. Biological Psychiatry 89, 627–636. doi: 10.1016/j.biopsych.2020.09.010.

Demontis, D., Walters, R. K., Martin, J., Mattheisen, M., Als, T. D., Agerbo, E., et al. (2019). Discovery of the first genome-wide significant risk loci for attention deficit/hyperactivity disorder. Nat Genet 51, 63–75. doi: 10.1038/s41588-018-0269-7.

Edwards, S. L., Beesley, J., French, J. D., and Dunning, A. M. (2013). Beyond GWASs: Illuminating the Dark Road from Association to Function. The American Journal of Human Genetics 93, 779–797. doi: 10.1016/j.ajhg.2013.10.012.

Ferkingstad, E., Sulem, P., Atlason, B. A., Sveinbjornsson, G., Magnusson, M. I., Styrmisdottir, E. L., et al. (2021). Large-scale integration of the plasma proteome with genetics and disease. Nature Genetics 53, 1712–1721. doi: 10.1038/s41588-021-00978-w.

Firth, J., Teasdale, S. B., Allott, K., Siskind, D., Marx, W., Cotter, J., et al. (2019). The efficacy and safety of nutrient supplements in the treatment of mental disorders: a meta-review of meta-analyses of randomized controlled trials. World Psychiatry 18, 308–324. doi: 10.1002/wps.20672.

Francis, M., Sun, Y., Xu, H., Brenna, J. T., and Ye, K. (2022). Fifty-one novel and replicated GWAS loci for polyunsaturated and monounsaturated fatty acids in 124,024 Europeans. 2022.05.27.22275343. doi: 10.1101/2022.05.27.22275343.

Gamazon, E. R., Wheeler, H. E., Shah, K. P., Mozaffari, S. V., Aquino-Michaels, K., Carroll, R. J., et al. (2015). A gene-based association method for mapping traits using reference transcriptome data. Nat Genet 47, 1091–1098. doi: 10.1038/ng.3367.

Gelernter, J., Kranzler, H. R., Sherva, R., Almasy, L., Koesterer, R., Smith, A. H., et al. (2014). Genome-wide association study of alcohol dependence:significant findings in African- and European-Americans including novel risk loci. Mol Psychiatry 19, 41–49. doi: 10.1038/mp.2013.145.

Gelernter, J., Sun, N., Polimanti, R., Pietrzak, R., Levey, D. F., Bryois, J., et al. (2019). Genome-wide association study of post-traumatic stress disorder reexperiencing symptoms in >165,000 US veterans. Nat Neurosci 22, 1394–1401. doi: 10.1038/s41593-019-0447-7.

Gleason, K. J., Yang, F., Pierce, B. L., He, X., and Chen, L. S. (2020). Primo: integration of multiple GWAS and omics QTL summary statistics for elucidation of molecular mechanisms of trait-associated SNPs and detection of pleiotropy in complex traits. Genome Biol 21, 1–24. doi: 10.1186/s13059-020-02125-w.

Gold, L., Ayers, D., Bertino, J., Bock, C., Bock, A., Brody, E. N., et al. (2010). Aptamer-Based Multiplexed Proteomic Technology for Biomarker Discovery. PLOS ONE 5, e15004. doi: 10.1371/journal.pone.0015004.

Gold, L., Walker, J. J., Wilcox, S. K., and Williams, S. (2012). Advances in human proteomics at high scale with the SOMAscan proteomics platform. New Biotechnology 29, 543–549. doi: 10.1016/j.nbt.2011.11.016.

Grove, J., Ripke, S., Als, T. D., Mattheisen, M., Walters, R. K., Won, H., et al. (2019). Identification of common genetic risk variants for autism spectrum disorder. Nat Genet 51, 431–444. doi: 10.1038/s41588-019-0344-8.

Gusev, A., Ko, A., Shi, H., Bhatia, G., Chung, W., Penninx, B. W. J. H., et al. (2016). Integrative approaches for large-scale transcriptome-wide association studies. Nat Genet 48, 245–252. doi: 10.1038/ng.3506.

Hammerschlag, A. R., Byrne, E. M., Agbessi, M., Ahsan, H., Alves, I., Andiappan, A., et al. (2020). Refining Attention-Deficit/Hyperactivity Disorder and Autism Spectrum Disorder Genetic Loci by Integrating Summary Data From Genome-wide Association, Gene Expression, and DNA Methylation Studies. Biological Psychiatry 88, 470–479. doi: 10.1016/j.biopsych.2020.05.002.

Hannon, E., Dempster, E., Viana, J., Burrage, J., Smith, A. R., Macdonald, R., et al. (2016). An integrated genetic-epigenetic analysis of schizophrenia: evidence for co-localization of genetic associations and differential DNA methylation. Genome Biology 17, 176. doi: 10.1186/s13059-016-1041-x.

Hatoum, A. S., Colbert, S. M. C., Johnson, E. C., Huggett, S. B., Deak, J. D., Pathak, G., et al. (2022). Multivariate genome-wide association meta-analysis of over 1 million subjects identifies loci underlying multiple substance use disorders. medRxiv, 2022.01.06.22268753. doi: 10.1101/2022.01.06.22268753.

Howard, D. M., Adams, M. J., Clarke, T.-K., Hafferty, J. D., Gibson, J., Shirali, M., et al. (2019). Genome-wide meta-analysis of depression identifies 102 independent variants and highlights the importance of the prefrontal brain regions. Nat Neurosci 22, 343–352. doi: 10.1038/s41593-018-0326-7.

Howard, D. M., Pain, O., Arathimos, R., Barbu, M. C., Amador, C., Walker, R. M., et al. (2022). Methylome-wide association study of early life stressors and adult mental health. Human Molecular Genetics 31, 651–664. doi: 10.1093/hmg/ddab274.

Hu, Y., Li, M., Lu, Q., Weng, H., Wang, J., Zekavat, S. M., et al. (2019). A statistical framework for cross-tissue transcriptome-wide association analysis. Nat Genet 51, 568–576. doi: 10.1038/s41588-019-0345-7.

Jaffe, A. E., Gao, Y., Deep-Soboslay, A., Tao, R., Hyde, T. M., Weinberger, D. R., et al. (2016). Mapping DNA methylation across development, genotype and schizophrenia in the human frontal cortex. Nat Neurosci 19, 40–47. doi: 10.1038/nn.4181.

Johnson, E. C., Demontis, D., Thorgeirsson, T. E., Walters, R. K., Polimanti, R., Hatoum, A. S., et al. (2020). A large-scale genome-wide association study meta-analysis of cannabis use disorder. The Lancet Psychiatry 7, 1032–1045. doi: 10.1016/S2215-0366(20)30339-4.

Johnson, E. C., Kapoor, M., Hatoum, A. S., Zhou, H., Polimanti, R., Wendt, F. R., et al. (2021). Investigation of convergent and divergent genetic influences underlying schizophrenia and alcohol use disorder. Psychological Medicine, 1–9. doi: 10.1017/S003329172100266X.

Kapoor, M., Chao, M. J., Johnson, E. C., Novikova, G., Lai, D., Meyers, J. L., et al. (2021). Multi-omics integration analysis identifies novel genes for alcoholism with potential overlap with neurodegenerative diseases. Nat Commun 12, 5071. doi: 10.1038/s41467-021-25392-y.

Kelley, K. W., Nakao-Inoue, H., Molofsky, A. V., and Oldham, M. C. (2018). Variation among intact tissue samples reveals the core transcriptional features of human CNS cell classes. Nat Neurosci 21, 1171–1184. doi: 10.1038/s41593-018-0216-z.

Kranzler, H. R., Zhou, H., Kember, R. L., Vickers Smith, R., Justice, A. C., Damrauer, S., et al. (2019). Genome-wide association study of alcohol consumption and use disorder in 274,424 individuals from multiple populations. Nat Commun 10, 1499. doi: 10.1038/s41467-019-09480-8.

Lee, P. H., Anttila, V., Won, H., Feng, Y.-C. A., Rosenthal, J., Zhu, Z., et al. (2019). Genomic Relationships, Novel Loci, and Pleiotropic Mechanisms across Eight Psychiatric Disorders. Cell 179, 1469-1482.e11. doi: 10.1016/j.cell.2019.11.020.

Liao, C., Laporte, A. D., Spiegelman, D., Akçimen, F., Joober, R., Dion, P. A., et al. (2019). Transcriptome-wide association study of attention deficit hyperactivity disorder identifies associated genes and phenotypes. Nat Commun 10, 4450. doi: 10.1038/s41467-019-12450-9.

Liu, Y., Chen, S., Li, Z., Morrison, A. C., Boerwinkle, E., and Lin, X. (2019). ACAT: A Fast and Powerful p Value Combination Method for Rare-Variant Analysis in Sequencing Studies. The American Journal of Human Genetics 104, 410–421. doi: 10.1016/j.ajhg.2019.01.002.

Mancuso, N., Freund, M. K., Johnson, R., Shi, H., Kichaev, G., Gusev, A., et al. (2019). Probabilistic fine-mapping of transcriptome-wide association studies. Nat Genet 51, 675–682. doi: 10.1038/s41588-019-0367-1.

Marsh, W. K., Penny, J. L., and Rothschild, A. J. (2017). Vitamin D supplementation in bipolar depression: A double blind placebo controlled trial. Journal of Psychiatric Research 95, 48–53. doi: 10.1016/j.jpsychires.2017.07.021.

Marstrand, T. T., and Storey, J. D. (2014). Identifying and mapping cell-type-specific chromatin programming of gene expression. Proceedings of the National Academy of Sciences 111, E645–E654. doi: 10.1073/pnas.1312523111.

McRae, A. F., Marioni, R. E., Shah, S., Yang, J., Powell, J. E., Harris, S. E., et al. (2018). Identification of 55,000 Replicated DNA Methylation QTL. Sci Rep 8, 17605. doi: 10.1038/s41598-018-35871-w.

Mocking, R. J. T., Harmsen, I., Assies, J., Koeter, M. W. J., Ruhé, H. G., and Schene, A. H. (2016). Meta-analysis and meta-regression of omega-3 polyunsaturated fatty acid supplementation for major depressive disorder. Transl Psychiatry 6, e756–e756. doi: 10.1038/tp.2016.29.

Mullins, N., Forstner, A. J., O’Connell, K. S., Coombes, B., Coleman, J. R. I., Qiao, Z., et al. (2021). Genome-wide association study of more than 40,000 bipolar disorder cases provides new insights into the underlying biology. Nat Genet 53, 817–829. doi: 10.1038/s41588-021-00857-4.

Nagpal, S., Meng, X., Epstein, M. P., Tsoi, L. C., Patrick, M., Gibson, G., et al. (2019). TIGAR: An Improved Bayesian Tool for Transcriptomic Data Imputation Enhances Gene Mapping of Complex Traits. The American Journal of Human Genetics 105, 258–266. doi: 10.1016/j.ajhg.2019.05.018.

Ng, B., White, C. C., Klein, H.-U., Sieberts, S. K., McCabe, C., Patrick, E., et al. (2017). An xQTL map integrates the genetic architecture of the human brain’s transcriptome and epigenome. Nat Neurosci 20, 1418–1426. doi: 10.1038/nn.4632.

Nievergelt, C. M., Maihofer, A. X., Klengel, T., Atkinson, E. G., Chen, C.-Y., Choi, K. W., et al. (2019). International meta-analysis of PTSD genome-wide association studies identifies sex- and ancestry-specific genetic risk loci. Nat Commun 10, 4558. doi: 10.1038/s41467-019-12576-w.

Niu, H.-M., Yang, P., Chen, H.-H., Hao, R.-H., Dong, S.-S., Yao, S., et al. (2019). Comprehensive functional annotation of susceptibility SNPs prioritized 10 genes for schizophrenia. Transl Psychiatry 9, 1–12. doi: 10.1038/s41398-019-0398-5.

Okereke, O. I., Reynolds, C. F., Iii, Mischoulon, D., Chang, G., Vyas, C. M., Cook, N. R., et al. (2020). Effect of Long-term Vitamin D3 Supplementation vs Placebo on Risk of Depression or Clinically Relevant Depressive Symptoms and on Change in Mood Scores: A Randomized Clinical Trial. JAMA 324, 471–480. doi: 10.1001/jama.2020.10224.

Ongen, H., Brown, A. A., Delaneau, O., Panousis, N. I., Nica, A. C., and Dermitzakis, E. T. (2017). Estimating the causal tissues for complex traits and diseases. Nat Genet 49, 1676–1683. doi: 10.1038/ng.3981.

Owen, M. J., and Williams, N. M. (2021). Explaining the missing heritability of psychiatric disorders. World Psychiatry 20, 294–295. doi: 10.1002/wps.20870.

Pardiñas, A. F., Holmans, P., Pocklington, A. J., Escott-Price, V., Ripke, S., Carrera, N., et al. (2018). Common schizophrenia alleles are enriched in mutation-intolerant genes and in regions under strong background selection. Nat Genet 50, 381–389. doi: 10.1038/s41588-018-0059-2.

Pathak, G. A., Singh, K., Wendt, F. R., Fleming, T. W., Overstreet, C., Koller, D., et al. (2022). Genetically regulated multi-omics study for symptom clusters of posttraumatic stress disorder highlights pleiotropy with hematologic and cardio-metabolic traits. Mol Psychiatry 27, 1394–1404. doi: 10.1038/s41380-022-01488-9.

Perzel Mandell, K. A., Eagles, N. J., Wilton, R., Price, A. J., Semick, S. A., Collado-Torres, L., et al. (2021). Genome-wide sequencing-based identification of methylation quantitative trait loci and their role in schizophrenia risk. Nat Commun 12, 5251. doi: 10.1038/s41467-021-25517-3.

Plana-Ripoll, O., Pedersen, C. B., Holtz, Y., Benros, M. E., Dalsgaard, S., de Jonge, P., et al. (2019). Exploring Comorbidity Within Mental Disorders Among a Danish National Population. JAMA Psychiatry 76, 259–270. doi: 10.1001/jamapsychiatry.2018.3658.

Polimanti, R., Walters, R. K., Johnson, E. C., McClintick, J. N., Adkins, A. E., Adkins, D. E., et al. (2020). Leveraging genome-wide data to investigate differences between opioid use vs. opioid dependence in 41,176 individuals from the Psychiatric Genomics Consortium. Mol Psychiatry 25, 1673–1687. doi: 10.1038/s41380-020-0677-9.

Qi, T., Wu, Y., Fang, H., Zhang, F., Liu, S., Zeng, J., et al. (2022). Genetic control of RNA splicing and its distinct role in complex trait variation. Nat Genet, 1–9. doi: 10.1038/s41588-022-01154-4.

Qi, T., Wu, Y., Zeng, J., Zhang, F., Xue, A., Jiang, L., et al. (2018). Identifying gene targets for brain-related traits using transcriptomic and methylomic data from blood. Nat Commun 9, 2282. doi: 10.1038/s41467-018-04558-1.

Reay, W. R., and Cairns, M. J. (2020). Pairwise common variant meta-analyses of schizophrenia with other psychiatric disorders reveals shared and distinct gene and gene-set associations. Transl Psychiatry 10, 134. doi: 10.1038/s41398-020-0817-7.

Robins, C., Liu, Y., Fan, W., Duong, D. M., Meigs, J., Harerimana, N. V., et al. (2021). Genetic control of the human brain proteome. The American Journal of Human Genetics 108, 400–410. doi: 10.1016/j.ajhg.2021.01.012.

Sanchez-Priego, C., Hu, R., Boshans, L. L., Lalli, M., Janas, J. A., Williams, S. E., et al. (2022). Mapping cis-regulatory elements in human neurons links psychiatric disease heritability and activity-regulated transcriptional programs. Cell Reports 39, 110877. doi: 10.1016/j.celrep.2022.110877.

Sanchez-Roige, S., Palmer, A. A., Fontanillas, P., Elson, S. L., 23 andMe Research Team, the Substance Use Disorder Working Group of the Psychiatric Genomics Consortium, Adams, M. J., et al. (2019). Genome-Wide Association Study Meta-Analysis of the Alcohol Use Disorders Identification Test (AUDIT) in Two Population-Based Cohorts. Am J Psychiatry 176, 107–118. doi: 10.1176/appi.ajp.2018.18040369.

Sarris, J., Murphy, J., Mischoulon, D., Papakostas, G. I., Fava, M., Berk, M., et al. (2016). Adjunctive Nutraceuticals for Depression: A Systematic Review and Meta-Analyses. AJP 173, 575–587. doi: 10.1176/appi.ajp.2016.15091228.

Scarr, E., Udawela, M., and Dean, B. (2019). Changed cortical risk gene expression in major depression and shared changes in cortical gene expression between major depression and bipolar disorders. Aust N Z J Psychiatry 53, 1189–1198. doi: 10.1177/0004867419857808.

Schrode, N., Ho, S.-M., Yamamuro, K., Dobbyn, A., Huckins, L., Matos, M. R., et al. (2019). Synergistic effects of common schizophrenia risk variants. Nature Genetics 51, 1475–1485. doi: 10.1038/s41588-019-0497-5.

Sekar, A., Bialas, A. R., de Rivera, H., Davis, A., Hammond, T. R., Kamitaki, N., et al. (2016). Schizophrenia risk from complex variation of complement component 4. Nature 530, 177–183. doi: 10.1038/nature16549.

Shen, X., Caramaschi, D., Adams, M. J., Walker, R. M., Min, J. L., Kwong, A., et al. (2022). DNA methylome-wide association study of genetic risk for depression implicates antigen processing and immune responses. Genome Medicine 14, 36. doi: 10.1186/s13073-022-01039-5.

Spaethling, J. M., Na, Y.-J., Lee, J., Ulyanova, A. V., Baltuch, G. H., Bell, T. J., et al. (2017). Primary Cell Culture of Live Neurosurgically Resected Aged Adult Human Brain Cells and Single Cell Transcriptomics. Cell Reports 18, 791–803. doi: 10.1016/j.celrep.2016.12.066.

Stein, M. B., Chen, C.-Y., Jain, S., Jensen, K. P., He, F., Heeringa, S. G., et al. (2017). Genetic risk variants for social anxiety. American Journal of Medical Genetics Part B: Neuropsychiatric Genetics 174, 120–131. doi: 10.1002/ajmg.b.32520.

Sugawara, H., Murata, Y., Ikegame, T., Sawamura, R., Shimanaga, S., Takeoka, Y., et al. (2018). DNA methylation analyses of the candidate genes identified by a methylome-wide association study revealed common epigenetic alterations in schizophrenia and bipolar disorder. Psychiatry and Clinical Neurosciences 72, 245–254. doi: 10.1111/pcn.12645.

Sun, B. B., Maranville, J. C., Peters, J. E., Stacey, D., Staley, J. R., Blackshaw, J., et al. (2018). Genomic atlas of the human plasma proteome. Nature 558, 73–79. doi: 10.1038/s41586-018-0175-2.

Taraszka, K., Zaitlen, N., and Eskin, E. (2022). Leveraging pleiotropy for joint analysis of genome-wide association studies with per trait interpretations. PLOS Genetics 18, e1010447. doi: 10.1371/journal.pgen.1010447.

THE BRAINSTORM CONSORTIUM, Anttila, V., Bulik-Sullivan, B., Finucane, H. K., Walters, R. K., Bras, J., et al. (2018). Analysis of shared heritability in common disorders of the brain. Science 360, eaap8757. doi: 10.1126/science.aap8757.

THE GTEX CONSORTIUM, Ardlie, K. G., Deluca, D. S., Segrè, A. V., Sullivan, T. J., Young, T. R., et al. (2015). The Genotype-Tissue Expression (GTEx) pilot analysis: Multitissue gene regulation in humans. Science 348, 648–660. doi: 10.1126/science.1262110.

Trubetskoy, V., Pardiñas, A. F., Qi, T., Panagiotaropoulou, G., Awasthi, S., Bigdeli, T. B., et al. (2022). Mapping genomic loci implicates genes and synaptic biology in schizophrenia. Nature 604, 502–508. doi: 10.1038/s41586-022-04434-5.

Turley, P., Walters, R. K., Maghzian, O., Okbay, A., Lee, J. J., Fontana, M. A., et al. (2018). Multi-trait analysis of genome-wide association summary statistics using MTAG. Nat Genet 50, 229–237. doi: 10.1038/s41588-017-0009-4.

van der Wijst, M., de Vries, D., Groot, H., Trynka, G., Hon, C., Bonder, M., et al. (2020). The single-cell eQTLGen consortium. eLife 9, e52155. doi: 10.7554/eLife.52155.

Võsa, U., Claringbould, A., Westra, H.-J., Bonder, M. J., Deelen, P., Zeng, B., et al. (2021). Large-scale cis- and trans-eQTL analyses identify thousands of genetic loci and polygenic scores that regulate blood gene expression. Nat Genet 53, 1300–1310. doi: 10.1038/s41588-021-00913-z.

Wainberg, M., Sinnott-Armstrong, N., Mancuso, N., Barbeira, A. N., Knowles, D. A., Golan, D., et al. (2019). Opportunities and challenges for transcriptome-wide association studies. Nat Genet 51, 592–599. doi: 10.1038/s41588-019-0385-z.

Wang, D., Liu, S., Warrell, J., Won, H., Shi, X., Navarro, F. C. P., et al. (2018). Comprehensive functional genomic resource and integrative model for the human brain. Science 362, eaat8464. doi: 10.1126/science.aat8464.

Watanabe, K., Taskesen, E., van Bochoven, A., and Posthuma, D. (2017). Functional mapping and annotation of genetic associations with FUMA. Nat Commun 8, 1826. doi: 10.1038/s41467-017-01261-5.

Watson, H. J., Yilmaz, Z., Thornton, L. M., Hübel, C., Coleman, J. R. I., Gaspar, H. A., et al. (2019). Genome-wide association study identifies eight risk loci and implicates metabo-psychiatric origins for anorexia nervosa. Nat Genet 51, 1207–1214. doi: 10.1038/s41588-019-0439-2.

Wingo, T. S., Liu, Y., Gerasimov, E. S., Gockley, J., Logsdon, B. A., Duong, D. M., et al. (2021). Brain proteome-wide association study implicates novel proteins in depression pathogenesis. Nat Neurosci 24, 810–817. doi: 10.1038/s41593-021-00832-6.

Wingo, T. S., Liu, Y., Gerasimov, E. S., Vattathil, S. M., Wynne, M. E., Liu, J., et al. (2022). Shared mechanisms across the major psychiatric and neurodegenerative diseases. Nat Commun 13, 4314. doi: 10.1038/s41467-022-31873-5.

Wu, Y., Zeng, J., Zhang, F., Zhu, Z., Qi, T., Zheng, Z., et al. (2018). Integrative analysis of omics summary data reveals putative mechanisms underlying complex traits. Nat Commun 9, 918. doi: 10.1038/s41467-018-03371-0.

Yang, C., Farias, F. H. G., Ibanez, L., Suhy, A., Sadler, B., Fernandez, M. V., et al. (2021a). Genomic atlas of the proteome from brain, CSF and plasma prioritizes proteins implicated in neurological disorders. Nat Neurosci 24, 1302–1312. doi: 10.1038/s41593-021-00886-6.

Yang, H., Liu, D., Zhao, C., Feng, B., Lu, W., Yang, X., et al. (2021b). Mendelian randomization integrating GWAS and eQTL data revealed genes pleiotropically associated with major depressive disorder. Translational Psychiatry 11, 225. doi: 10.1038/s41398-021-01348-0.

Yuan, Z., Zhu, H., Zeng, P., Yang, S., Sun, S., Yang, C., et al. (2020). Testing and controlling for horizontal pleiotropy with probabilistic Mendelian randomization in transcriptome-wide association studies. Nat Commun 11, 3861. doi: 10.1038/s41467-020-17668-6.

Zhang, J., Dutta, D., Köttgen, A., Tin, A., Schlosser, P., Grams, M. E., et al. (2021). Large Bi-Ethnic Study of Plasma Proteome Leads to Comprehensive Mapping of cis-pQTL and Models for Proteome-wide Association Studies. doi: 10.1101/2021.03.15.435533.

Zhang, S., Zhang, H., Zhou, Y., Qiao, M., Zhao, S., Kozlova, A., et al. (2020). Allele-specific open chromatin in human iPSC neurons elucidates functional disease variants. Science 369, 561–565. doi: 10.1126/science.aay3983.

Zhou, D., Jiang, Y., Zhong, X., Cox, N. J., Liu, C., and Gamazon, E. R. (2020). A unified framework for joint-tissue transcriptome-wide association and Mendelian randomization analysis. Nat Genet 52, 1239–1246. doi: 10.1038/s41588-020-0706-2.

Zhu, Z., Zhang, F., Hu, H., Bakshi, A., Robinson, M. R., Powell, J. E., et al. (2016). Integration of summary data from GWAS and eQTL studies predicts complex trait gene targets. Nat Genet 48, 481–487. doi: 10.1038/ng.3538.

