## Supplementary material for "Identifying potential risk genes and pathways for neuropsychiatric and substance use disorders using intermediate molecular mediator information"

### 1 Supplementary Method

#### 1.1 Reference molecular xQTL mapping data

To increase power, large consortia often meta-analyzed many smaller xQTL mapping studies. Consequently, to increase discovery power, we often use such large e/p/m-QTL studies as reference data for our XWAS analyses. When allele frequency information was missing in these reference data, we substituted the allele frequencies from European cohort from the 1000 Genome phase 3 [1] (503 individuals) (https://ctg.cncr.nl/software/MAGMA/ref_data/g1000_eur.zip, accessed on August 3, 2022). We used the same reference panel as LD reference input for SMR. Below, we provide further details about the xQTL reference datasets.

#### Blood eQTL (eqtlGen)

This data set is comprised of meta-analysis of eQTL mapping from 37 different dataset. DNA isolation of the sample was obtained either from human whole-blood or peripheral blood mononuclear cells. For gene expression assay, they used Affymetrix expression array, Illumina expression array and RNA sequencing. At variant level, exclusion criteria were: MAF < 1%, HWE P value < $10^{-4}$ and MACH r2 score < 50%.

1.) A mapping method called SHRiMP aligner v2.2.3 [2] was used to align reads to genome build GRCh37/hg19. Probes mapping to multiple genomic regions were eliminated.
2.) To normalize the expression level measurements, quantile normalization was used. The expression data was log2 transformed. Based on these, outliers were excluded.
3.) To adjust the data for ancestral population stratification, the principal components were regressed out using Plink version 1.07 [3]. The model was adjusted for biological covariates and confounders. Then, residuals were used for the eQTL mapping. These steps were followed for Illumina gene expression array data.
4.) For Affymetrix data, the details were explained in the original article [4]. For RNA-seq data, extra steps are using trimmed mean of M-values normalization method [5] and inclusion criteria of having more than 50% count per million at minimum 1% of all samples for each probe.
5.) Another step of permutation testing (matching probes based on empirical test results) was conducted to form a shared set of genes between all studies.

Meta analysis was conducted by using optimally weighted Z-score method [6]. The cis-eQTL mapping method was the one used in Westra et al. [7]. Permutation based gene level FDR was used for multiple testing correction.

**Brain eQTL (BrainMeta version 2)**

This is a meta-analysis of the seven studies (sample size in parenthesis): BrainGVEX, Common Mind Consortium (CMC, n=469) [8], NIMH Human Brain Collection Core (HBCC, n=129), Lieber Institute for Brain Development (LIBD, n=128), Mount Sinai VA Medical Center Brain Bank (MSBB, n=635), Mayo Clinic (n=288), Religious Order Study and Rush Memory and Aging (ROS/MAP, n=832) [9]. There were 1,962,114 eQTL SNPs and 16,704 eGenes identified. ROS/MAP and MSBB used genotyping arrays and the remaining whole-genome sequencing. The meta-analysis was performed using MeCS method (https://yanglab.westlake.edu.cn/software/smr/#MeCS)[10].

1.) STAR v.2.7.8a [11]was used to align reads to genome build GRCh37/hg19.
2.) To quantify RNA abundance, they used RNA-SeQC v.2.3.5 [12].
3.) Exclusion criteria at transcript level was RNA integrity number below < 5.5.
4.) Each individual has to have more than 10 million transcripts read in total.
5.) At variant level, they used Plink version 2 to apply to exclude variants having a MAF < 1%, HWE P value < $10^{-6}$ and INFO score < 30%.

**Blood pQTL (deCODE)**

This a large scale pQTL mapping study based on 35,559 Icelandics. Study participants are from Icelandic Cancer project and deCODE genetics. The mean age of the cohort was 55. The proteins were isolated from blood plasma. For protein quantification, the authors used aptamer-based technology Slow Off-rate Modified Aptamer Scan (SOMAScan) [13,14] assay version 4. They included 4,907 aptamers that correspond to 4,719 proteins. Variants with MAF < 0.01 and INFO < 0.9 were excluded from protein GWAS.

The cis-eQTL mapping method was BOLT-LMM [15]. The significance threshold was set as $1.8 \times 10^{-9}$ (Bonferroni method adjusting for multiple testing). The definition for cis-pQTL: SNP with significant signal that is located within 1Mb of the transcription start site of the protein coding gene.

**Brain pQTL (Wingo et al.)**

Participants were recruited by ROS/MAP, Banner Sun Health Research Institute and MSSB studies. All participants are at later stage of life and mostly elderly individuals. They all donated organs to these institutions under an informed consent. Variants with MAF < 0.05, HWE P value < $5 \times 10^{-7}$ and genotype missing rate > 0.05 were excluded from protein GWAS. Only participants with European ancestry were included. Tissues were homogenized and proteins were digested before protein quantification step.

The quantification method used was isobaric tandem mass tag (TMT) labeling mass spectrophotometry. First, the proteins were labeled with Thermo Fisher Scientific kits (TMT 10 or 11-plex kits). Second, proteins were fractioned. Third step is the quantification with TMT mass spectrometry based on mass/charge ratio. Each study may have different protocols in these steps so, for more details, please see the original publication [16].

1.) Protein abundance measurements were log2 transformed.
2.) Possible confounders (batch, sex, age, etc.) were regressed out. Singular Value Decomposition was used to regress out effects of hidden confounders.
3.) Protein GWAS included 722 samples from all three studies and 9,363 proteins.

**Blood mQTL**

The mQTL reference data was generated from two major studies: Lothian Birth Cohorts (n=1,366)[17] and Brisbane Systems Genetics Study (n=614)[18]. The methylation assay was Illumina Human Methylation 450K. All samples were peripheral blood samples. They meta-analyzed these two studies by MeCS method.

**Brain mQTL**

This dataset is meta-analysis of three studies by Qi et al. [10] : ROS/MAP (n=468), Hanon et al. (n=166) and Jaffe et al. (526). Methylation assay is the same used in blood mQTL. The meta-analysis method is MeCS. At the end, results had 397,621 CpG methylation probes and approximately 7.7 million methylation SNPs.

**1.2 SMR XWAS analysis**

We performed blood and brain SMR XWAS analyses on GWAS summary statistics data from nine different major PDs. One the purpose of testing both blood and brain is finding out the concordance of the direction of probe level effect between blood and brain. The instrumental variable in SMR analysis is the top associated sentinel SNP for xQTL. Then, it uses genetic effect estimate on the phenotype ($\hat{\beta}_{GP}$) and on the exposure (gene expression/protein abundance/CpG probe) ($\hat{\beta}_{GE}$) to estimate the effect of exposure on the phenotype ($\hat{\beta}_{EP}$) by using the following formula: $\hat{\beta}_{EP} = \hat{\beta}_{GP}/\hat{\beta}_{GE}$ [19].

Trait GWAS summary statistics ($\hat{\beta}_{GP}$) and xQTL reference data ($\hat{\beta}_{GE}$) come from independent cohorts. For our analyses, we used only cis-xQTLs, i.e., SNPs with P value > $5x10^{-8}$ within the 2 Mb upstream and downstream of the probe. Positions for all variants and genes in input files (LD reference, GWAS summary statistics, and xQTL summary statistics files) for SMR analysis are based on GRCh37 reference genome.

### 2 Supplementary Figures

**2. Regional Manhattan plots for MHC region (chromosome 6)**

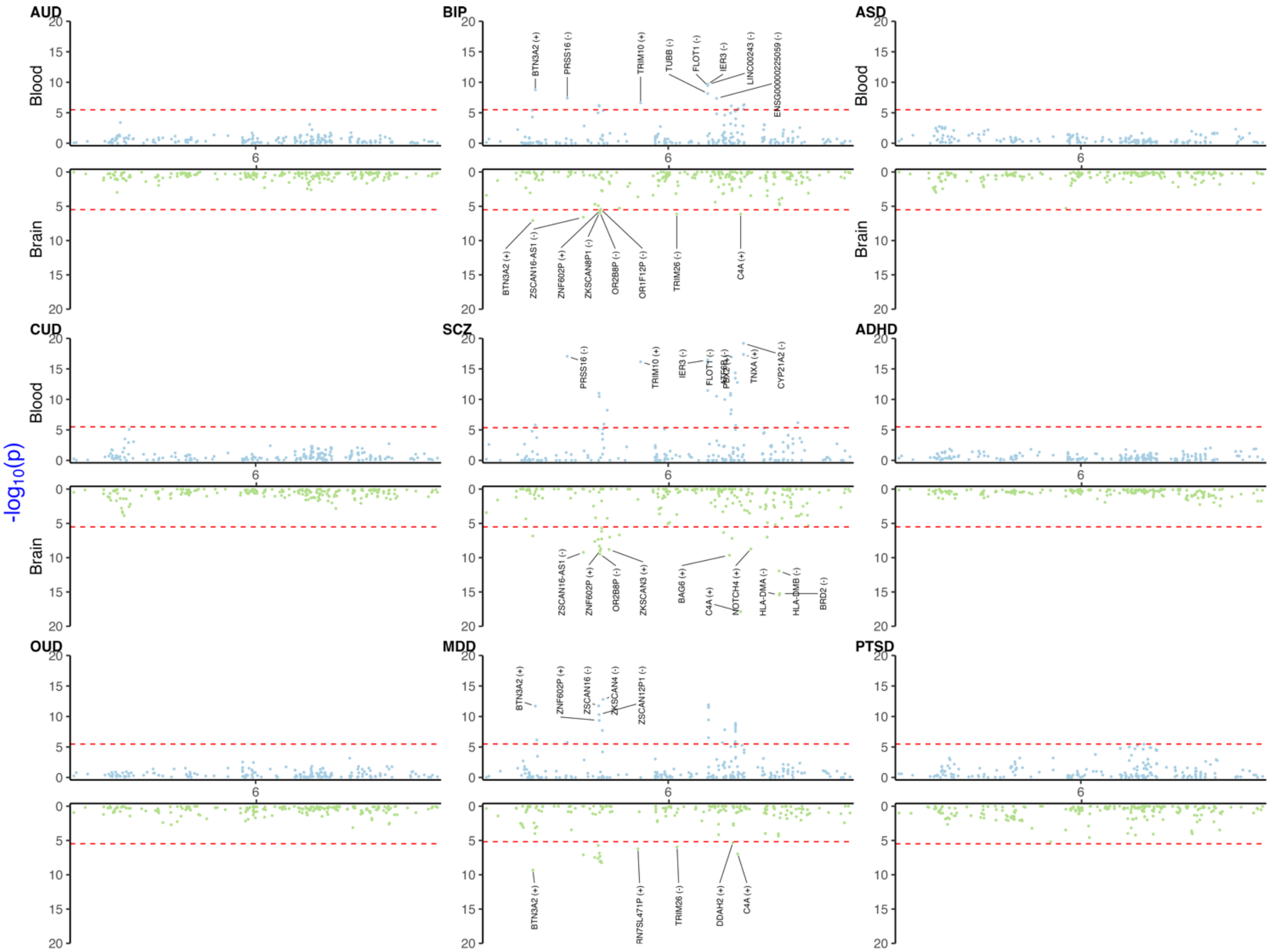


**Supplementary Figure 1.** TWAS Miami plot (Manhattan blood-brain bi-plot) for MHC region.

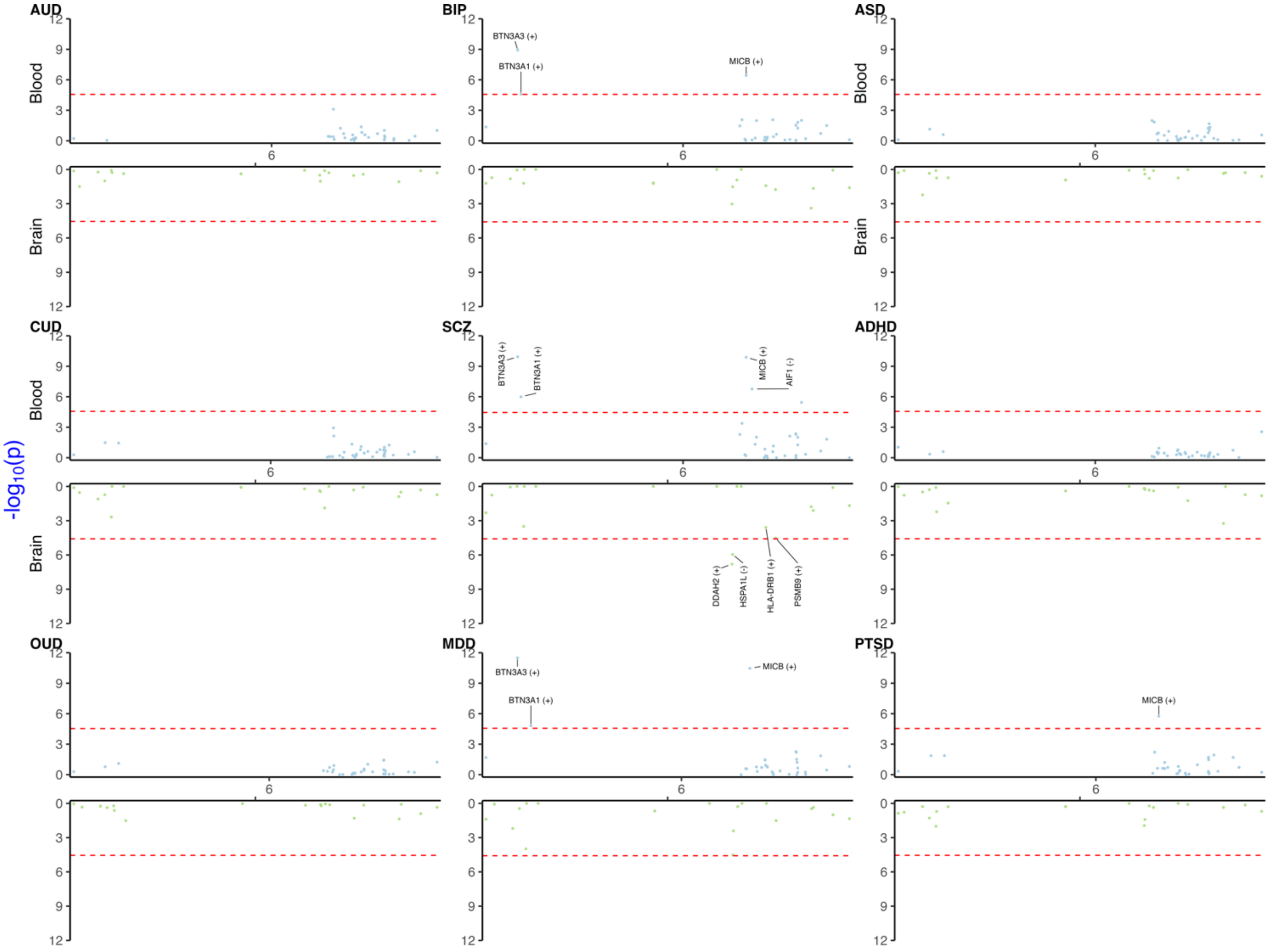


**Supplementary Figure 2.** PWAS Miami plot (Manhattan blood-brain bi-plot) for MHC region.

### 2.2 FUMA gene set enrichment results

We included all protein coding, processed transcripts, pseudogenes, long-noncoding RNA, and noncoding RNA genes to observe any significant enrichment. We selected Benjamini-Hochberg FDR procedure to adjust gene set enrichment P value for multiple testing. All the FUMA gene-to-function gene set enrichment analyses excluded the genes in MHC region.

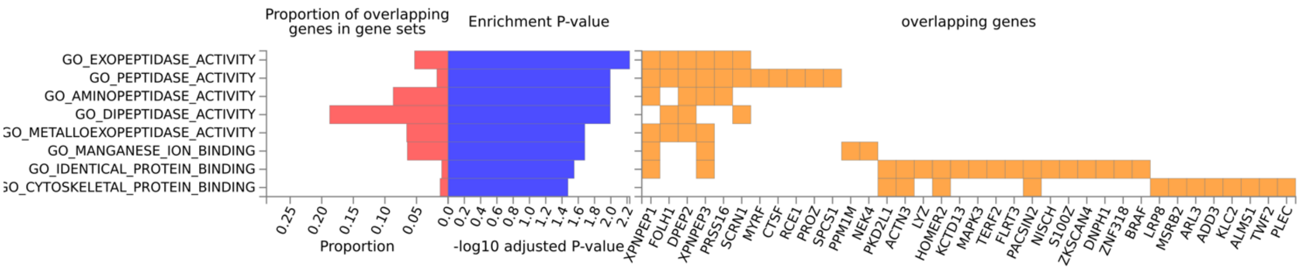


**Supplementary Figure 3.** Go Molecular Function gene set enrichment results for BIP blood T/PWAS.

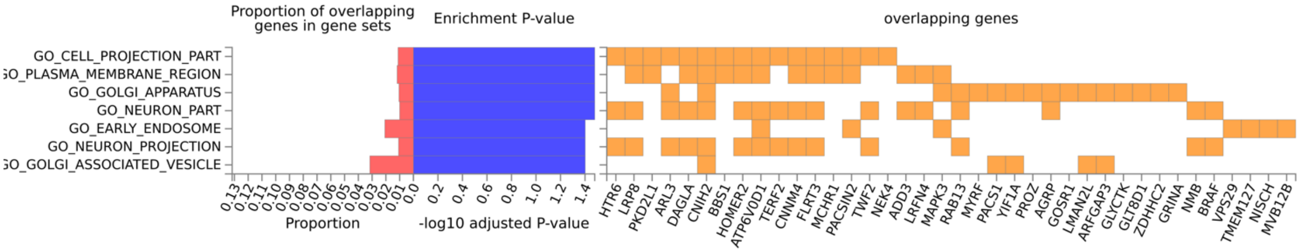


**Supplementary Figure 4.** Go Cellular Component gene set enrichment results for BIP blood T/PWAS.

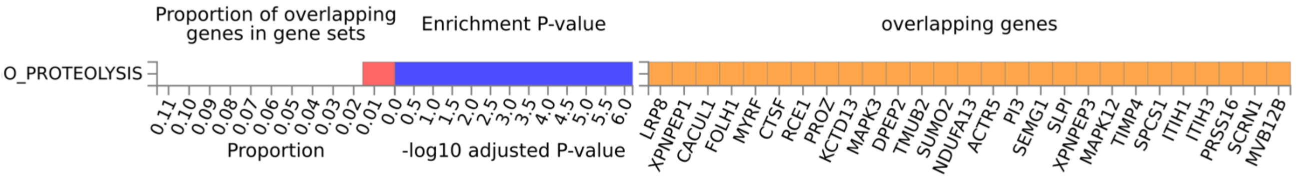


**Supplementary Figure 5.** Go Biological Process gene set enrichment results for BIP blood T/PWAS.

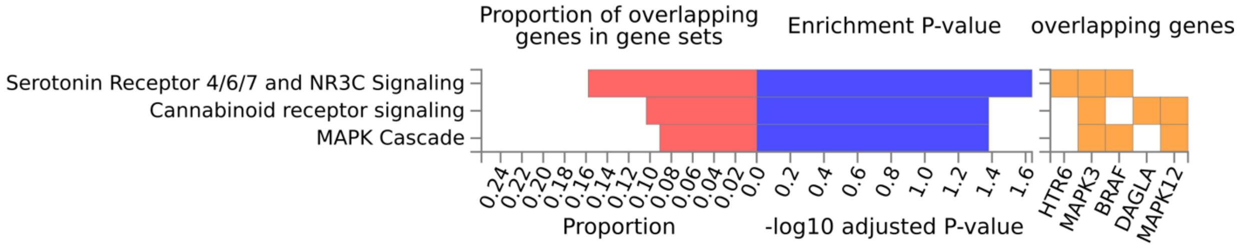


**Supplementary Figure 6.** Wiki Pathways gene set enrichment results for BIP blood T/PWAS.

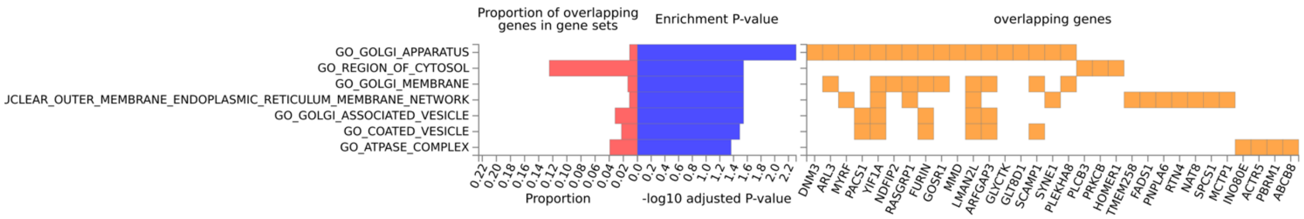


**Supplementary Figure 7.** Go Cellular Component gene set enrichment results for BIP brain T/PWAS.

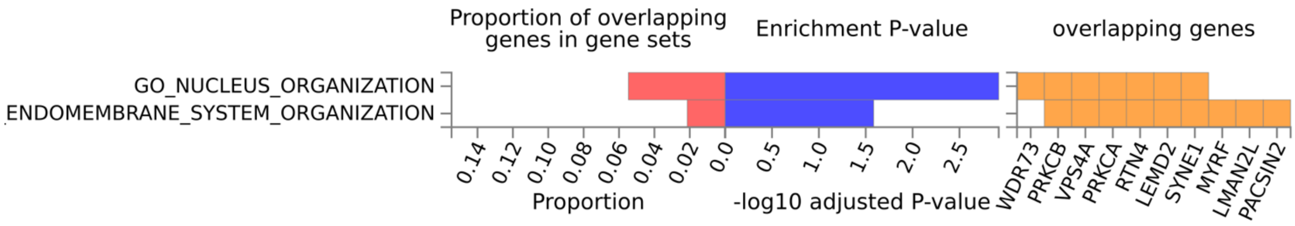


**Supplementary Figure 8.** Go Biological Process gene set enrichment results for BIP brain T/PWAS.

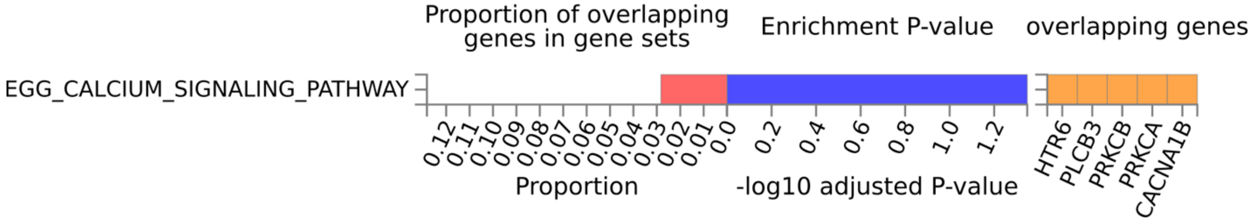


**Supplementary Figure 9.** KEGG gene set enrichment results for BIP brain T/PWAS.

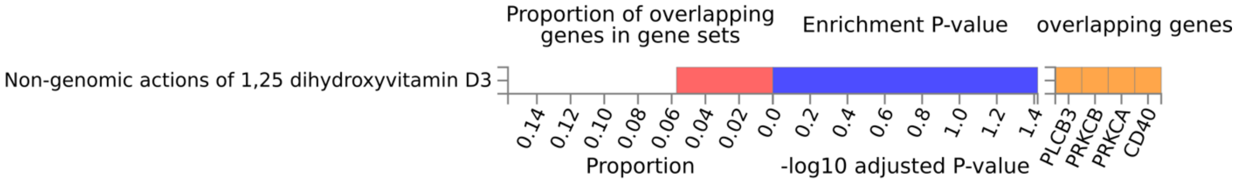


**Supplementary Figure 10.** Wiki Pathways gene set enrichment results for BIP brain T/PWAS.

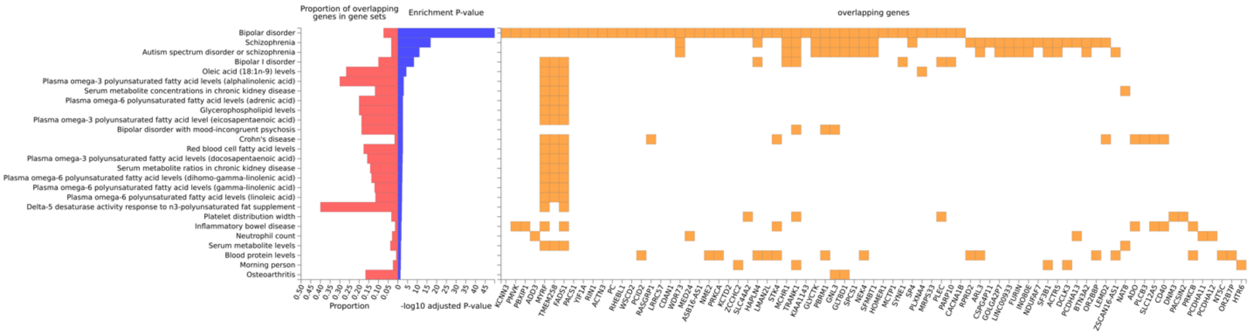


**Supplementary Figure 11.** GWAS catalog gene set enrichment results for BIP brain T/PWAS.

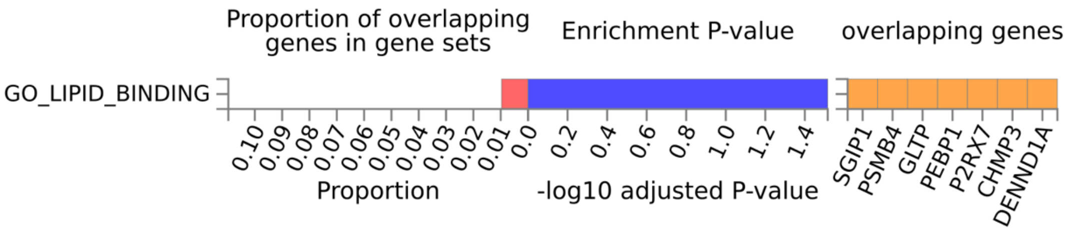


**Supplementary Figure 12.** Go Molecular Function gene set enrichment results for MDD brain T/PWAS

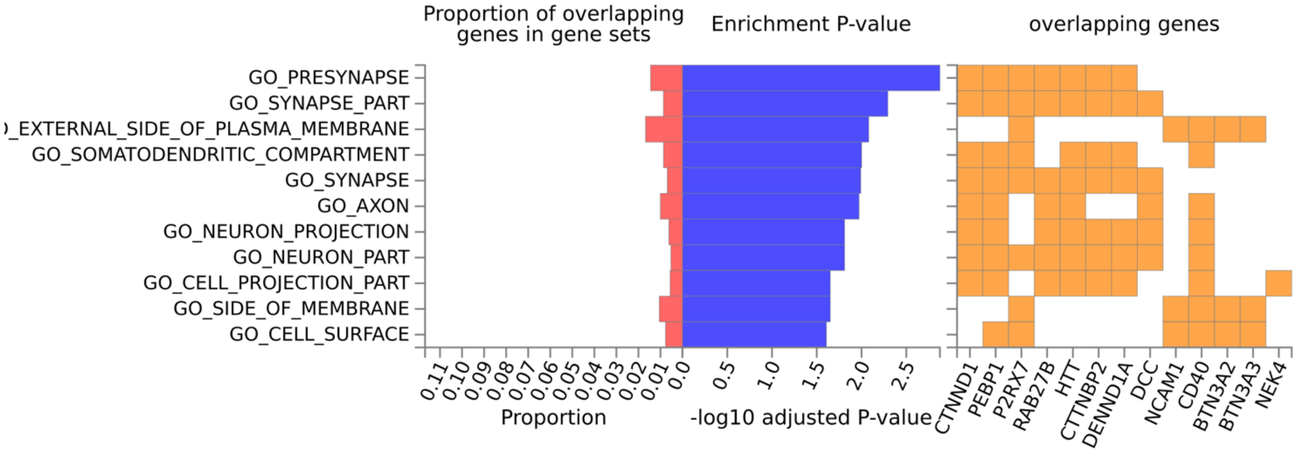


**Supplementary Figure 13.** Go Cellular Component gene set enrichment results for MDD brain T/PWAS.

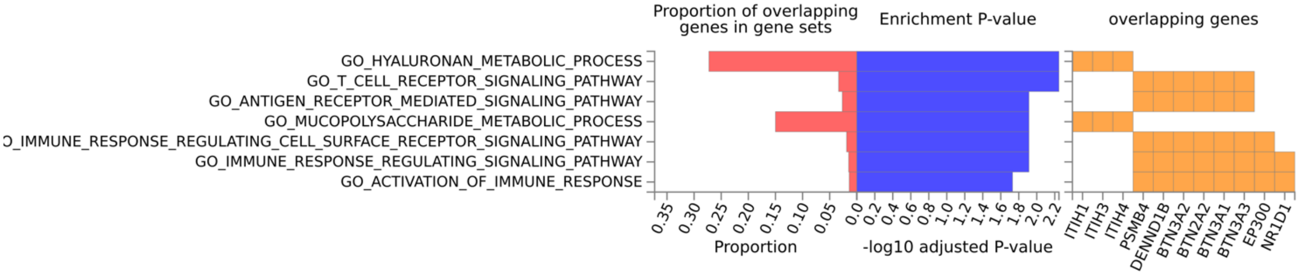


**Supplementary Figure 14.** Go Biological Process gene set enrichment results for MDD blood T/PWAS.

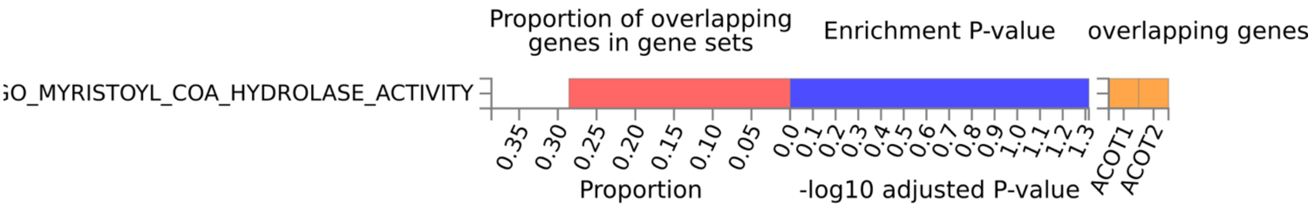


**Supplementary Figure 15.** Go Molecular Function gene set enrichment results for PTSD brain T/PWAS.

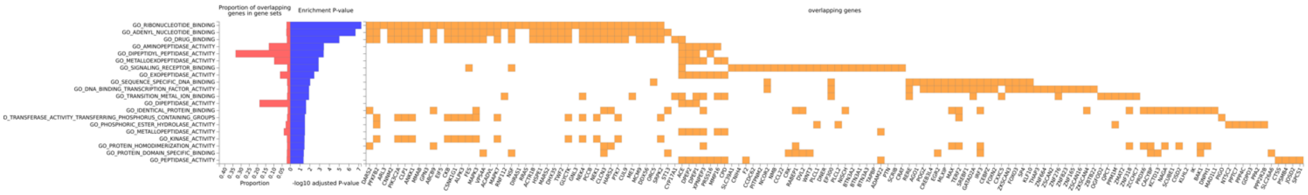


**Supplementary Figure 16.** Go Molecular Function gene set enrichment results for SCZ blood T/PWAS.

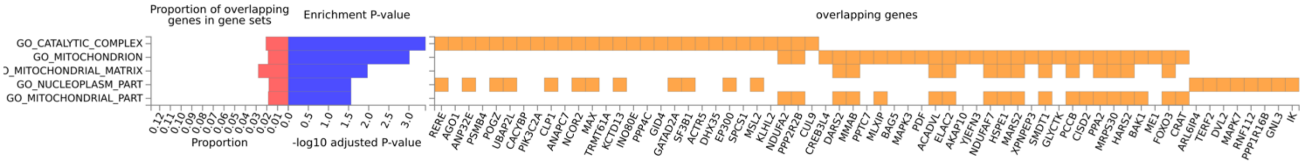


**Supplementary Figure 17.** Go Cellular Component gene set enrichment results for SCZ blood T/PWAS.

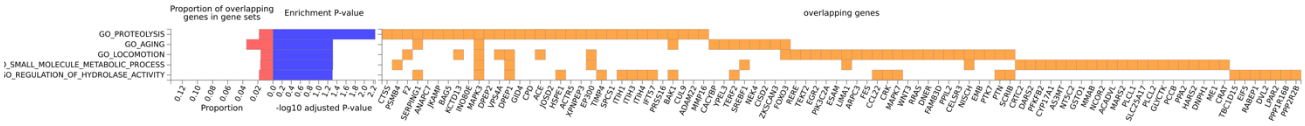


**Supplementary Figure 18.** Go Biological Process gene set enrichment results for SCZ blood T/PWAS.

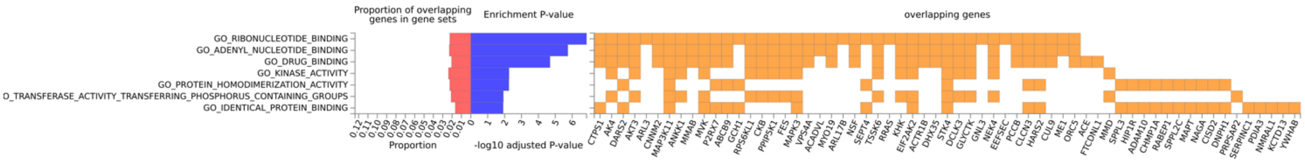


**Supplementary Figure 19.** Go Molecular Function gene set enrichment results for SCZ brain T/PWAS.

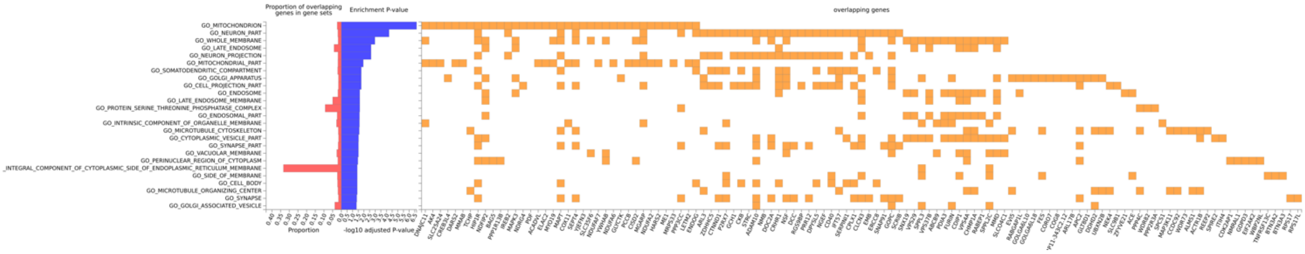


**Supplementary Figure 20.** Go Cellular Component gene set enrichment results for SCZ brain T/PWAS.

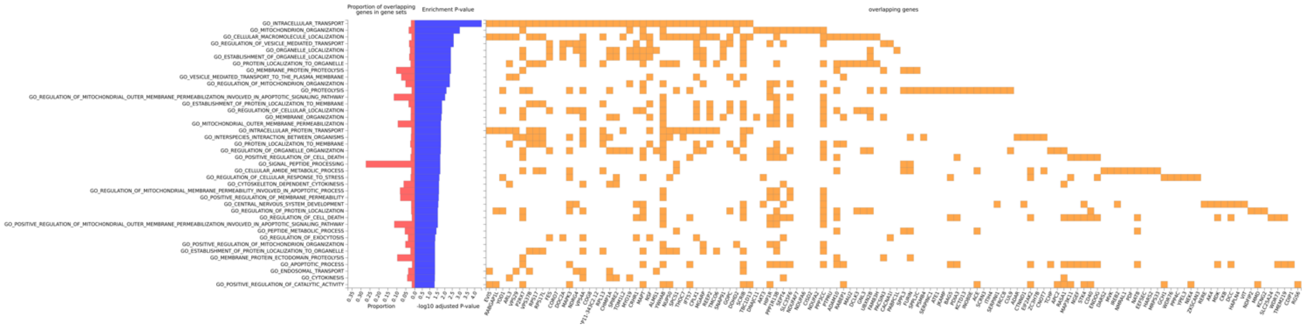


**Supplementary Figure 21.** Go Biological Process gene set enrichment results for SCZ brain T/PWAS.

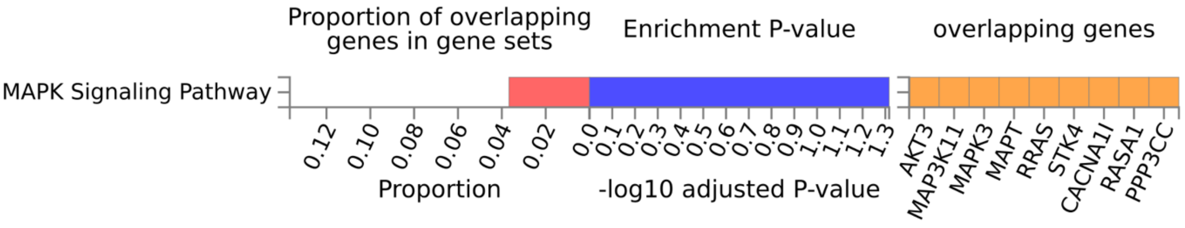


**Supplementary Figure 22.** Wiki Pathways gene set enrichment results for SCZ brain T/PWAS.

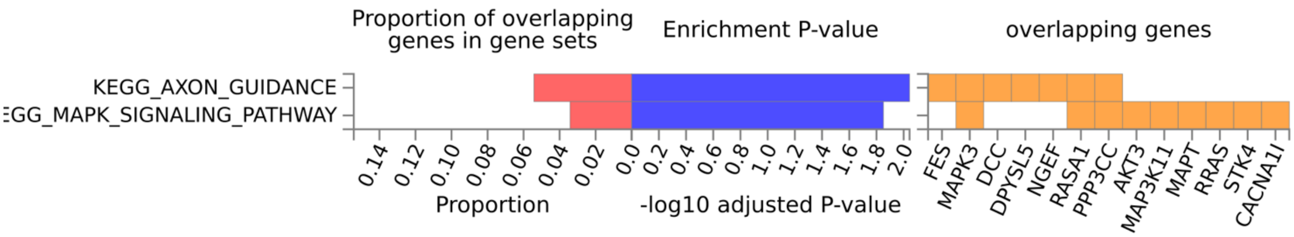


**Supplementary Figure 23.** KEGG gene set enrichment results for SCZ brain T/PWAS.

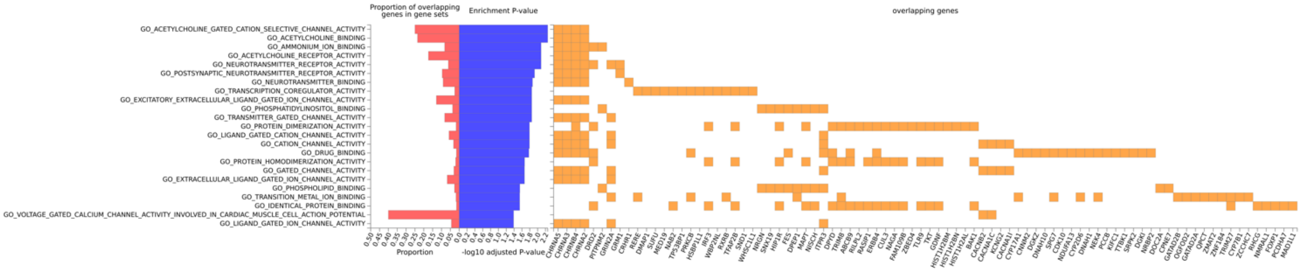


**Supplementary Figure 24.** Go Molecular Function gene set enrichment results for SCZ brain MWAS.

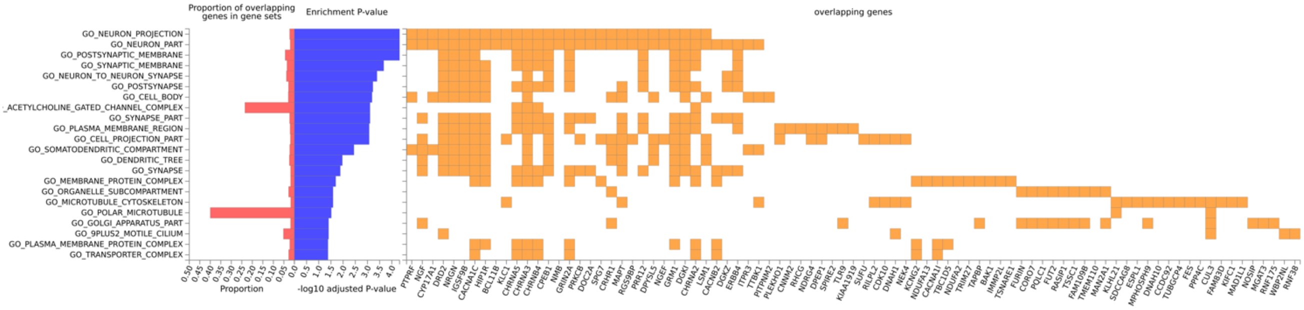


**Supplementary Figure 25.** Go Cellular Components gene set enrichment results for SCZ brain MWAS.

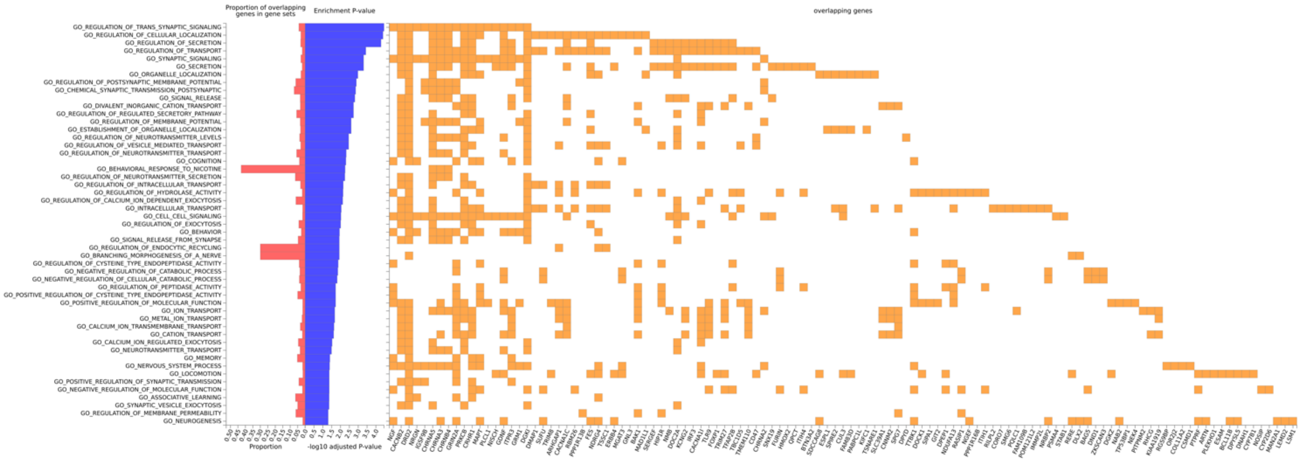

**Supplementary Figure 26.** Go Biological Process gene set enrichment results for SCZ brain MWAS.

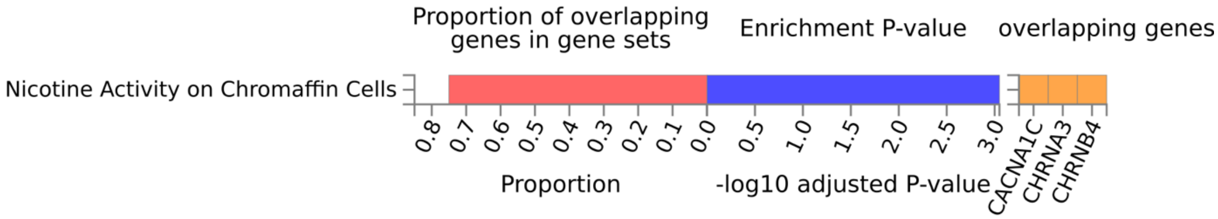


**Supplementary Figure 27.** Wiki Pathways gene set enrichment results for SCZ brain MWAS.

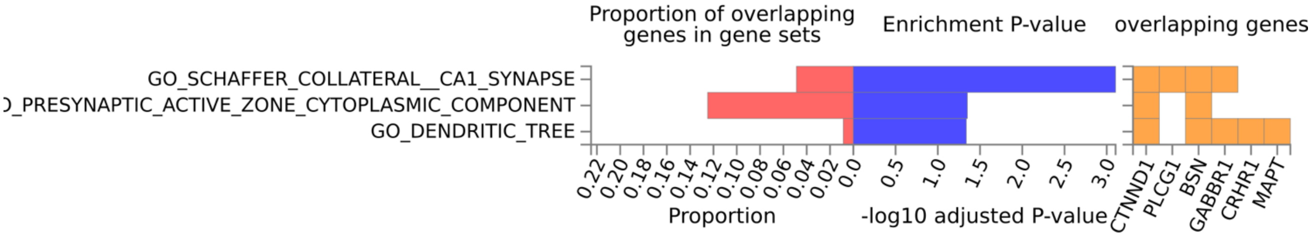


**Supplementary Figure 28.** Go Cellular Components gene set enrichment results for PTSD blood MWAS.

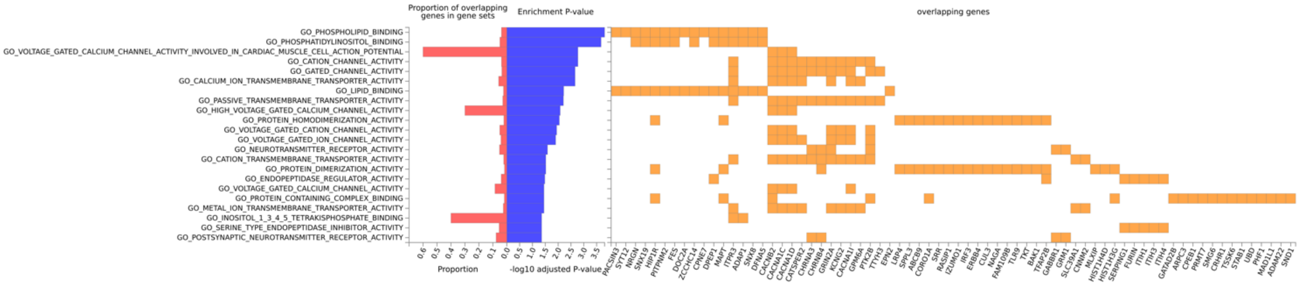

**Supplementary Figure 29.** Go Molecular Function gene set enrichment results for SCZ blood MWAS.

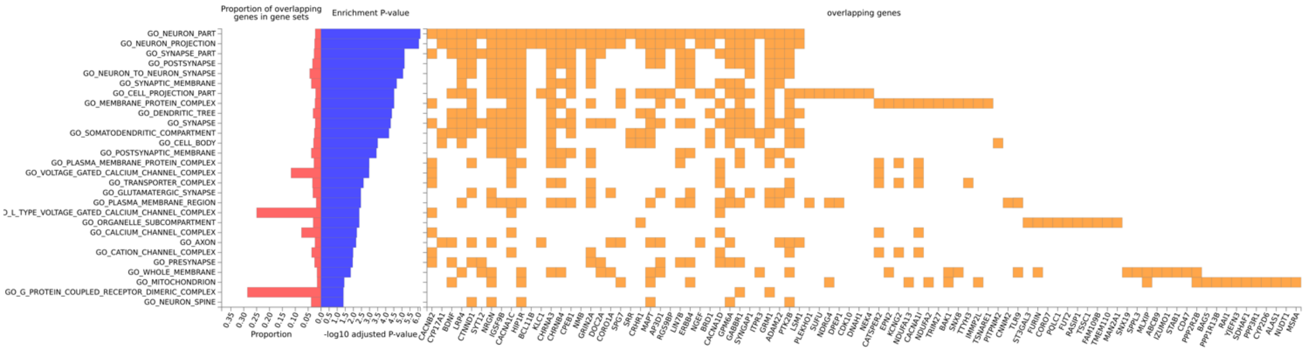


**Supplementary Figure 30.** Go Cellular Components FUMA gene set enrichment results for SCZ blood MWAS.

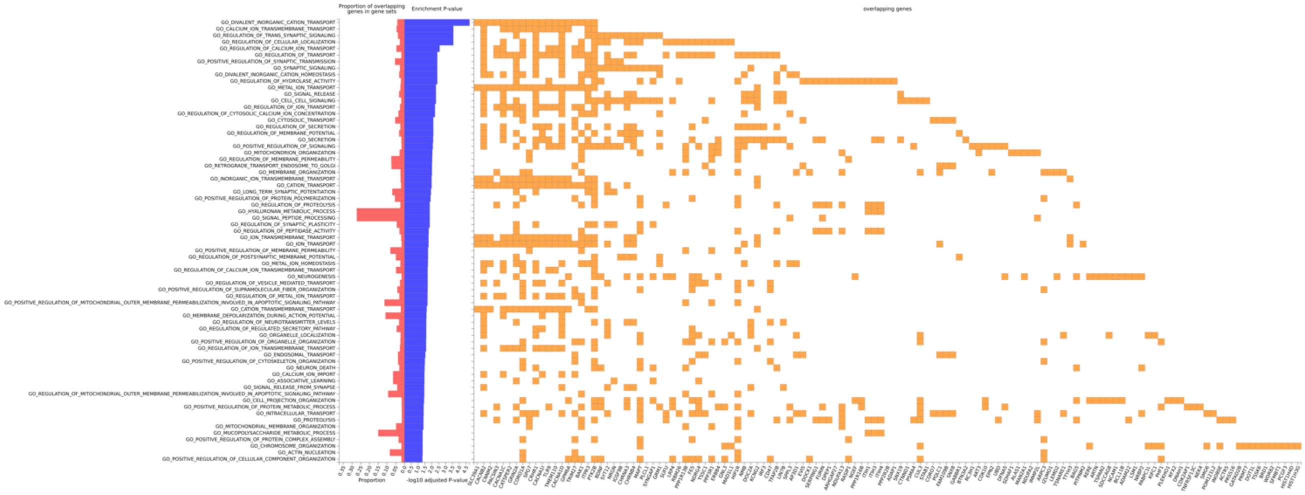

**Supplementary Figure 31.** Go Biological Process gene set enrichment results for SCZ blood MWAS.

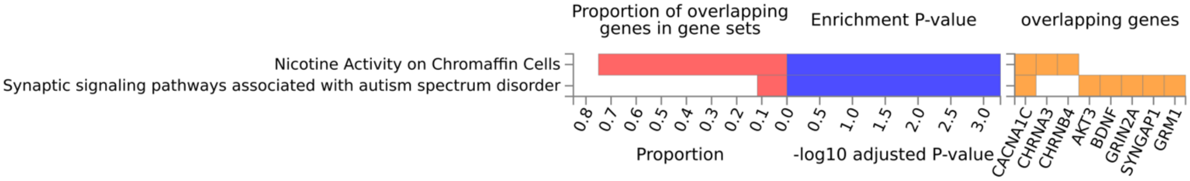


**Supplementary Figure 32.** Wiki Pathways gene set enrichment results for SCZ blood MWAS.

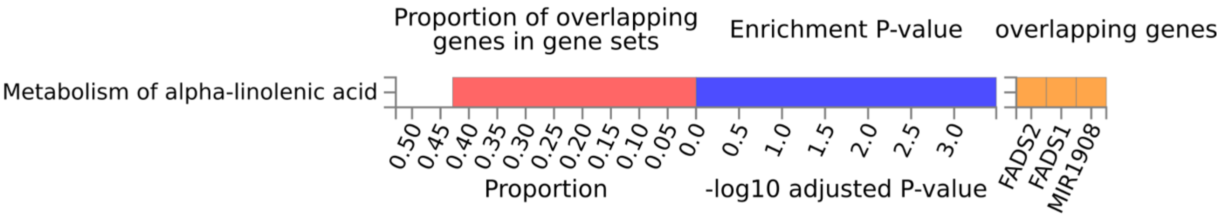

**Supplementary Figure 33.** Wiki Pathways gene set enrichment results for BIP blood MWAS.

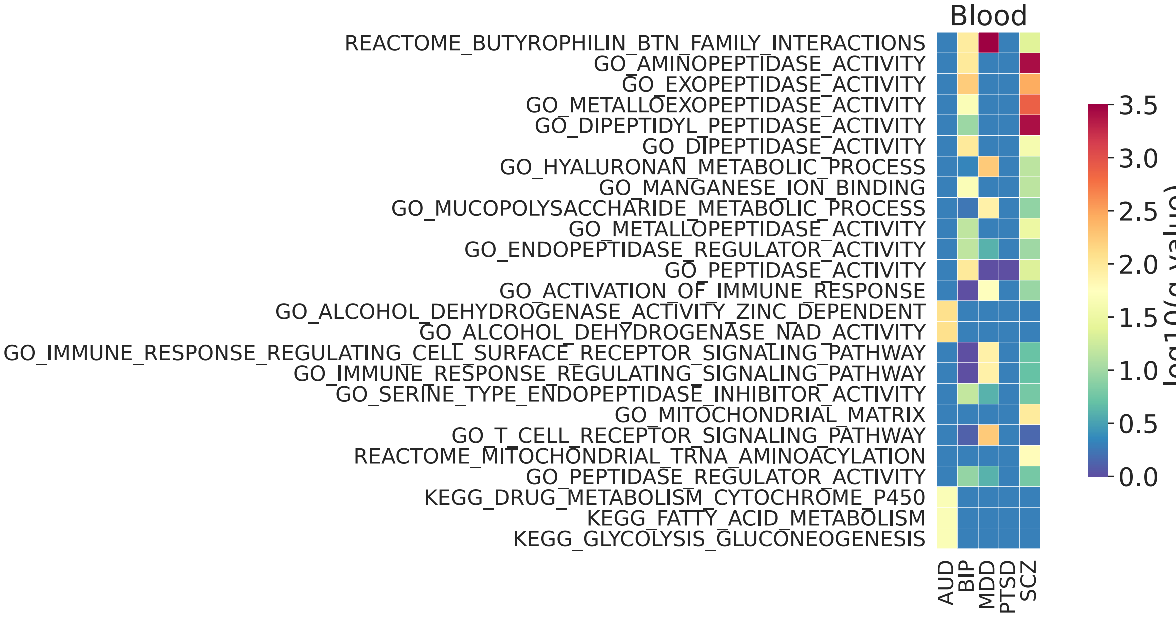


**Supplementary Figure.34** Gene set enrichment of combined TWAS and PWAS blood signals (genes with FDR q-value <0.05). Gene sets are ranked by the sum of -log10(q-value) across traits. (Including only GO term, Reactome and KEGG gene sets for simplicity)

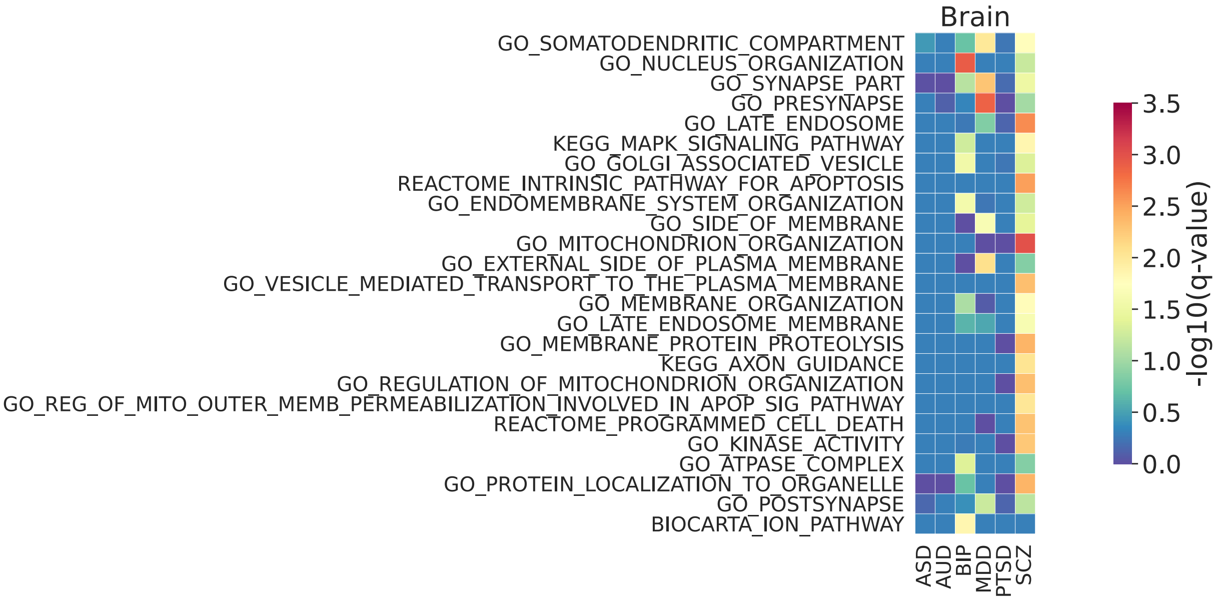


**Supplementary Figure.35** Gene set enrichment of combined TWAS and PWAS brain signals (genes with FDR q-value <0.05). Gene sets are ranked by the sum of -log10(q-value) across traits.

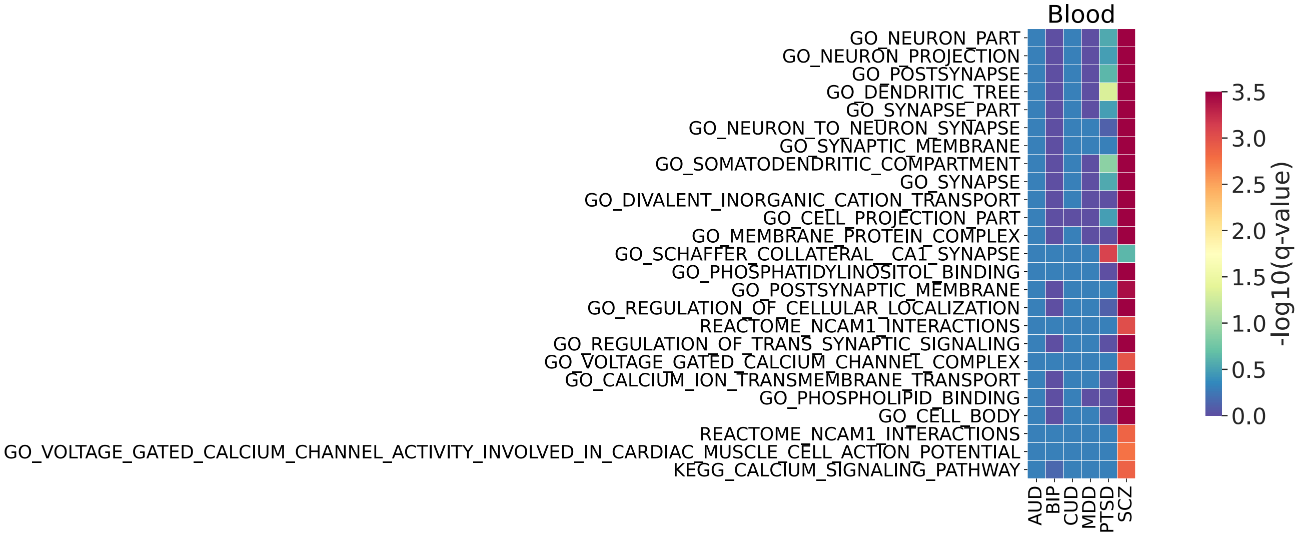


**Supplementary Figure.36** Gene set enrichment of MWAS only blood signals (genes with FDR q-value <0.05). Gene sets are ranked by the sum of -log10(q-value) across traits.

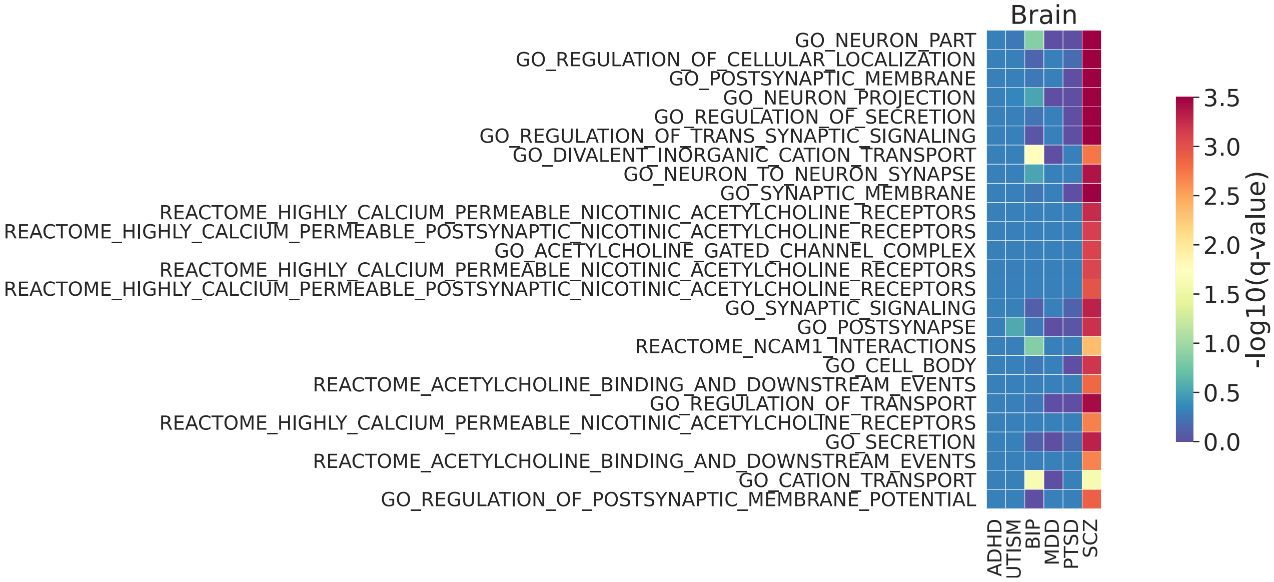


**Supplementary Figure.37** Gene set enrichment of MWAS only brain signals (genes with FDR q-value <0.05). Gene sets are ranked by the sum of -log10(q-value) across traits.

### References


1. Auton, A.; Abecasis, G.R.; Altshuler, D.M.; Durbin, R.M.; Abecasis, G.R.; Bentley, D.R.; Chakravarti, A.; Clark, A.G.; Donnelly, P.; Eichler, E.E.; et al. A Global Reference for Human Genetic Variation. *Nature* **2015**, *526*, 68–74, doi:10.1038/nature15393.

2. David, M.; Dzamba, M.; Lister, D.; Ilie, L.; Brudno, M. SHRiMP2: Sensitive yet Practical Short Read Mapping. *Bioinformatics* **2011**, *27*, 1011–1012, doi:10.1093/bioinformatics/btr046.

3. Purcell, S.; Neale, B.; Todd-Brown, K.; Thomas, L.; Ferreira, M.A.R.; Bender, D.; Maller, J.; Sklar, P.; de Bakker, P.I.W.; Daly, M.J.; et al. PLINK: A Tool Set for Whole-Genome Association and Population-Based Linkage Analyses. *Am J Hum Genet* **2007**, *81*, 559–575.

4. Võsa, U.; Claringbould, A.; Westra, H.-J.; Bonder, M.J.; Deelen, P.; Zeng, B.; Kirsten, H.; Saha, A.; Kreuzhuber, R.; Yazar, S.; et al. Large-Scale Cis- and Trans-EQTL Analyses Identify Thousands of Genetic Loci and Polygenic Scores That Regulate Blood Gene Expression. *Nat Genet* **2021**, *53*, 1300–1310, doi:10.1038/s41588-021-00913-z.

5. Robinson, M.D.; Oshlack, A. A Scaling Normalization Method for Differential Expression Analysis of RNA-Seq Data. *Genome Biology* **2010**, *11*, R25, doi:10.1186/gb-2010-11-3-r25.

6. Zaykin, D.V. Optimally Weighted Z-Test Is a Powerful Method for Combining Probabilities in Meta-Analysis. *Journal of Evolutionary Biology* **2011**, *24*, 1836–1841, doi:10.1111/j.1420-9101.2011.02297.x.

7. Westra, H.-J.; Peters, M.J.; Esko, T.; Yaghootkar, H.; Schurmann, C.; Kettunen, J.; Christiansen, M.W.; Fairfax, B.P.; Schramm, K.; Powell, J.E.; et al. Systematic Identification of Trans EQTLs as Putative Drivers of Known Disease Associations. *Nat Genet* **2013**, *45*, 1238–1243, doi:10.1038/ng.2756.

8. Fromer, M.; Roussos, P.; Sieberts, S.K.; Johnson, J.S.; Kavanagh, D.H.; Perumal, T.M.; Ruderfer, D.M.; Oh, E.C.; Topol, A.; Shah, H.R.; et al. Gene Expression Elucidates Functional Impact of Polygenic Risk for Schizophrenia. *Nature Neuroscience* **2016**, *19*, 1442–1453, doi:10.1038/nn.4399.

9. Ng, B.; White, C.C.; Klein, H.-U.; Sieberts, S.K.; McCabe, C.; Patrick, E.; Xu, J.; Yu, L.; Gaiteri, C.; Bennett, D.A.; et al. An XQTL Map Integrates the Genetic Architecture of the Human Brain's Transcriptome and Epigenome. *Nat Neurosci* **2017**, *20*, 1418–1426, doi:10.1038/nn.4632.

10. Qi, T.; Wu, Y.; Zeng, J.; Zhang, F.; Xue, A.; Jiang, L.; Zhu, Z.; Kemper, K.; Yengo, L.; Zheng, Z.; et al. Identifying Gene Targets for Brain-Related Traits Using Transcriptomic and Methylomic Data from Blood. *Nat Commun* **2018**, *9*, 2282, doi:10.1038/s41467-018-04558-1.

11. Dobin, A.; Davis, C.A.; Schlesinger, F.; Drenkow, J.; Zaleski, C.; Jha, S.; Batut, P.; Chaisson, M.; Gingeras, T.R. STAR: Ultrafast Universal RNA-Seq Aligner. *Bioinformatics* **2013**, *29*, 15–21, doi:10.1093/bioinformatics/bts635.

12. Graubert, A.; Aguet, F.; Ravi, A.; Ardlie, K.G.; Getz, G. RNA-SeQC 2: Efficient RNA-Seq Quality Control and Quantification for Large Cohorts. *Bioinformatics* **2021**, *37*, 3048–3050, doi:10.1093/bioinformatics/btab135.

13. Gold, L.; Ayers, D.; Bertino, J.; Bock, C.; Bock, A.; Brody, E.N.; Carter, J.; Dalby, A.B.; Eaton, B.E.; Fitzwater, T.; et al. Aptamer-Based Multiplexed Proteomic Technology for Biomarker Discovery. *PLOS ONE* **2010**, *5*, e15004, doi:10.1371/journal.pone.0015004.

14. Gold, L.; Walker, J.J.; Wilcox, S.K.; Williams, S. Advances in Human Proteomics at High Scale with the SOMAscan Proteomics Platform. *New Biotechnology* **2012**, *29*, 543–549, doi:10.1016/j.nbt.2011.11.016.

15. Loh, P.-R.; Tucker, G.; Bulik-Sullivan, B.K.; Vilhjálmsson, B.J.; Finucane, H.K.; Salem, R.M.; Chasman, D.I.; Ridker, P.M.; Neale, B.M.; Berger, B.; et al. Efficient Bayesian Mixed-Model Analysis Increases Association Power in Large Cohorts. *Nat Genet* **2015**, *47*, 284–290, doi:10.1038/ng.3190.

16. Wingo, T.S.; Liu, Y.; Gerasimov, E.S.; Vattathil, S.M.; Wynne, M.E.; Liu, J.; Lori, A.; Faundez, V.; Bennett, D.A.; Seyfried, N.T.; et al. Shared Mechanisms across the Major Psychiatric and Neurodegenerative Diseases. *Nat Commun* **2022**, *13*, 4314, doi:10.1038/s41467-022-31873-5.

17. Chen, B.H.; Marioni, R.E.; Colicino, E.; Peters, M.J.; Ward-Caviness, C.K.; Tsai, P.-C.; Roetker, N.S.; Just, A.C.; Demerath, E.W.; Guan, W.; et al. DNA Methylation-Based Measures of Biological Age: Meta-Analysis Predicting Time to Death. *Aging* **2016**, *8*, 1844–1865, doi:10.18632/aging.101020.

18. Powell, J.E.; Henders, A.K.; McRae, A.F.; Caracella, A.; Smith, S.; Wright, M.J.; Whitfield, J.B.; Dermitzakis, E.T.; Martin, N.G.; Visscher, P.M.; et al. The Brisbane Systems Genetics Study: Genetical Genomics Meets Complex Trait Genetics. *PLOS ONE* **2012**, *7*, e35430, doi:10.1371/journal.pone.0035430.

19. Wu, Y.; Zeng, J.; Zhang, F.; Zhu, Z.; Qi, T.; Zheng, Z.; Lloyd-Jones, L.R.; Marioni, R.E.; Martin, N.G.; Montgomery, G.W.; et al. Integrative Analysis of Omics Summary Data Reveals Putative Mechanisms Underlying Complex Traits. *Nat Commun* **2018**, *9*, 918, doi:10.1038/s41467-018-03371-0.
